# Monoclonal Antibody Treatment, Prophylaxis and Vaccines Combined to Reduce SARS CoV-2 Spread

**DOI:** 10.1101/2021.05.21.21257624

**Authors:** Mohamed A. Kamal, Andreas Kuznik, Luyuan Qi, Witold Więcek, Mohamed Hussein, Hazem E. Hassan, Kashyap Patel, Thomas Obadia, Masood Khaksar Toroghi, Daniela J. Conrado, Nidal Al-Huniti, Roman Casciano, Meagan P. O’Brien, Ruanne V. Barnabas, Myron S. Cohen, Patrick F. Smith

## Abstract

**Background:** Antiviral monoclonal antibodies (mAbs) developed for treatment of COVID-19 reduce the magnitude and duration of viral shedding and can thus potentially contribute to reducing transmission of the causative virus, severe acute respiratory coronavirus 2 (SARS-CoV-2). However, use of these mAbs in combination with a vaccine program has not been considered in public health strategic planning.

**Methods:** We developed an agent-based model to characterize SARS-CoV-2 transmission in the US population during an aggressive phase of the pandemic (October 2020 to April 2021), and simulated the effects on infections and mortality of combining mAbs as treatment and post-exposure prophylaxis (PEP) with a vaccine program plus non-pharmaceutical interventions. We also interrogated the impact of rapid diagnostic testing, increased mAb supply, and vaccine rollout.

**Findings:** Allocation of mAbs as PEP or targeting those ≥65 years provided the greatest incremental benefits relative to vaccine in averting infections and deaths, by up to 17% and 41%, respectively. Rapid testing, facilitating earlier diagnosis and mAb use, amplified these benefits. The model was sensitive to mAb supply; doubling supply further reduced infections and mortality, by up to two-fold, relative to vaccine. mAbs continued to provide incremental benefits even as proportion of the vaccinated population increased.

**Interpretation:** Use of anti-viral mAbs as treatment and PEP in combination with a vaccination program would substantially reduce SARS-CoV-2 transmission and pandemic burden. These results may help guide resource allocation and patient management decisions for COVID-19 and can also be used to inform public health policy for current and future pandemic preparedness.

**Funding:** Regeneron Pharmaceuticals.

## Introduction

It has been more than one year since COVID-19, caused by severe acute respiratory coronavirus 2 (SARS-CoV-2), was declared a global pandemic,^1^ with rapid spread of the virus, devastating effects on individuals and economies, and extensive implementation of non-pharmaceutical interventions (NPIs). There has also been success in developing diagnostic tests, vaccines, and new therapeutic modalities including monoclonal antibodies (mAbs) in efforts to treat COVID-19 and control transmission.

The extent to which NPIs can reduce SARS CoV-2 transmission is dependent on behavior changes and adherence.^2,3^ More recently, several vaccines have proven effective as pre-exposure prevention,and their deployment represents a long-term strategy to achieve herd immunity. Concurrently, mAbs against the SARS-CoV-2 spike protein were developed for both treatment and prevention of COVID-19. While vaccines require development of active immunity to COVID-19 over time, mAbs confer immediate passive immunity, providing protection to individuals with recent SARS-CoV-2 exposure.^4,5^ mAbs are anticipated to be effective independent of underlying host immune status, provided that they can neutralize variants of concern (VOC). mAbs may be useful as both pre-and post-exposure prophylaxis, especially for individuals exposed to COVID-19 who have not been vaccinated, cannot receive a vaccine, or fail to respond to a vaccine. The benefits of mAbs are likely to increase as logistical challenges associated with their clinical use are addressed. Moreover, mAbs may provide further benefit in reducing pandemic burden in countries where vaccination rates have been slow, and/or where highly transmissible VOC (still sensitive to drugs) have posed a challenge to global health, for example, the current situation in India.^6^

Monoclonal antibodies can also provide treatment for pre-symptomatic or symptomatic infections to prevent progression of COVID-19 to more severe phases.^7^ The US Food and Drug Administration (FDA) granted Emergency Use Authorization (EUA) to two mAb combinations for treatment of COVID-19 in non-hospitalized patients at high-risk of severe disease. One combination, developed by Eli Lilly, consists of bamlanivimab plus etesevimab,^8^ and the other, REGEN-COV that was developed by Regeneron Pharmaceuticals, consists of casirivimab and imdevimab.^9^

To date, use of mAbs to prevent COVID-19 transmission, in combination with a vaccine program, has not been considered in public health strategic planning. However, mAbs can contribute to reducing transmission and overall pandemic burden when used in a Treatment-as-Prevention (TasP) strategy, since they reduce the magnitude and duration of viral shedding in addition to reducing risk of COVID-19 progression in ambulatory patients.^7,10^ Furthermore, mAbs can serve as a bridge to immunity shortly after vaccination, or as an alternative to vaccination for people who cannot respond to a vaccine.

Multiple COVID-19 prevention strategies are ongoing concomitantly, thus it is difficult to generate real-time, controlled, clinical trial data to understand their combined benefits. Mathematical modeling is an important tool to inform decision-making in scenarios where prospective evaluation of pandemic interventions in concert may not be feasible. Our objective is to apply such modeling to explore the role that mAbs may play in combination with vaccines to reduce the pandemic burden.

We developed an agent-based model (ABM) to characterize the individual-level heterogeneity of SARS-CoV-2 transmission in the US population. This model enabled simulation of various pandemic scenarios, relative to a base case that assumed implementation of an aggregate of NPIs. These scenarios included vaccine and mAbs in combination with vaccine. The mAbs were used as active treatment for TasP, and as passive immunity for post-exposure prophylaxis (PEP). We also assessed rapid diagnostic testing paired with mAb interventions as an additional intervention for increasing the effectiveness of TasP.

## Methods

Agent-based models can simulate behaviors of individuals (called “agents”), and interactions among them, to better characterize transmission dynamics of infectious diseases in a large population. An ABM was considered appropriate for our study, since it is flexible enough to simulate scenarios that assess virus transmission and the impact of different mitigation strategies across the US population. The schematic of the ABM (figure 1) shows linkage among the component modules that include population structure, social contact networks, agent movement or travel, virus transmission, disease progression, and interventions. Complete methodology, including data sources and model assumptions, is provided in the appendix.

**Figure 1.**
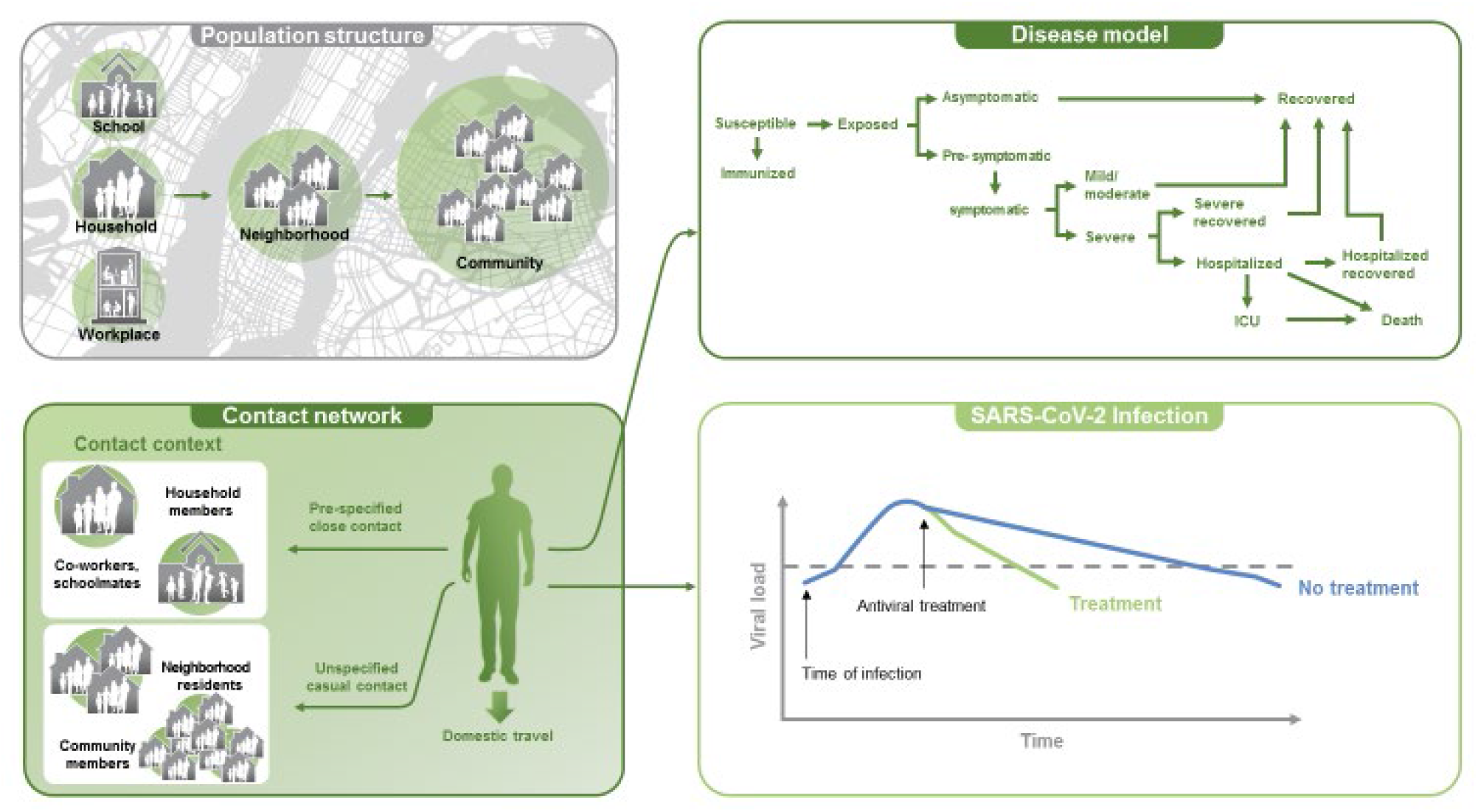
Components of the COVID-19 agent-based model. Development of the agent-based model incorporated 7 modules to allow for simulation of mitigation strategies for the COVID-19 pandemic. Components of the model consisted of the US population structure, a base social network, movement/travel within the US, virus transmission, a disease model, a baseline use of non-pharmaceutical interventions, and pharmaceutical interventions.

### US population structure

Agents are simulated to resemble the US population in terms of demographics (age and sex), household composition, group living settings (nursing home and student housing), education, and employment status. Agents are mapped based on population density to different locations across the US using individual-level information from the most recent 5-year aggregated data from US Census Bureau (appendix p 20).

### Social contact network

Since SARS-CoV-2 can be transmitted by pre-symptomatic and asymptomatic individuals^11,12^ with substantial secondary infection risk in households,^13,14^ modeling of social contact networks is crucial to characterizing transmission (appendix p 2). These networks consist of household, workplace, school, neighborhood, and community, representing different “mixing groups” of individuals (figure 1). The household, school, and workplace represent transmission pathways, with household as the primary pathway; communities and neighborhoods reflect occasional casual contacts.

### Movement and travel

Since our simulation scenarios were from late October 2020 onwards, we focused on propagation of COVID-19 inside the US, assuming the risk from imported cases would be negligible. Data on US domestic travel by air and car, including number of trips taken, purpose, and destinations, were derived from the US Department of Transportation and the US Travel Association (appendix p 22). Since trip length varies, we accounted for duration of population mixing in the new location.

### Virus transmission

The daily probability that a susceptible individual (S) is infected, P(t), depends on: age of the S; risk from imported cases; and risk of infection in mixing groups of S. We calculate this probability using the formula shown in Equation 1, which is based on person-to-person transmission.^15,16^

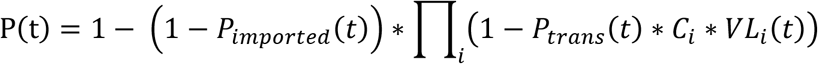

**Equation 1**. Formula for calculating the probability of being infected at each simulation time step

2*P*_*imported*_(*t*) ∗ *C*_*i*_ ∗ *VL*_*i*_(*t*) represents the probability for S to be infected by one contacted individual *i* in the same mixing groups; *C*_*i*_ represents the probability of a sufficient contact for transmission during one time step, dependent on the age and the mixing groups of S. *P*_*imported*_(t) is a scalar used to adjust the transmission probabilities to obtain the desired longitudinal mortality curve for target simulation scenarios. *VL*_*i*_(*t*) represents infectiousness of the agent, assumed to be proportional to the log10 of the daily viral load of that agent.^17^ *P*_*imported*_(*t*) represents the probability that S is infected by an imported case, which was assumed to be negligible.

### Disease progression

Each agent has a defined COVID-19 related health status (figure 1) that is updated in the disease model at each simulation time step (1 day). In the model, an exposed susceptible agent who acquires an infection can follow different disease progression pathways leading to recovery or death based on age-adjusted probabilities. We differentiated between pre-symptomatic and symptomatic individuals and between mild/moderate and severe disease so that the natural history of COVID–19 infections is realistically described and reflects how transmission forces change during the disease course (appendix p 4).

### Model calibration

To determine how well the model represents the real-world, the ABM was calibrated using US mortality data including longitudinal daily deaths reported by the Institute for Health Metrics Evaluation (IHME) and infection fatality ratio from the CDC (appendix p 28). The calibration spanned the period October 26, 2020 to April 4, 2021 (figure 2), which was considered an aggressive phase of the pandemic because of high mortality and overburdened health care systems, and during which time vaccines became available. The observed data show that mortality peaked from late October 2020 through January 2021. The calibrated model tracked closely to the actual number of deaths over time and cumulative mortality.

**Figure 2.**
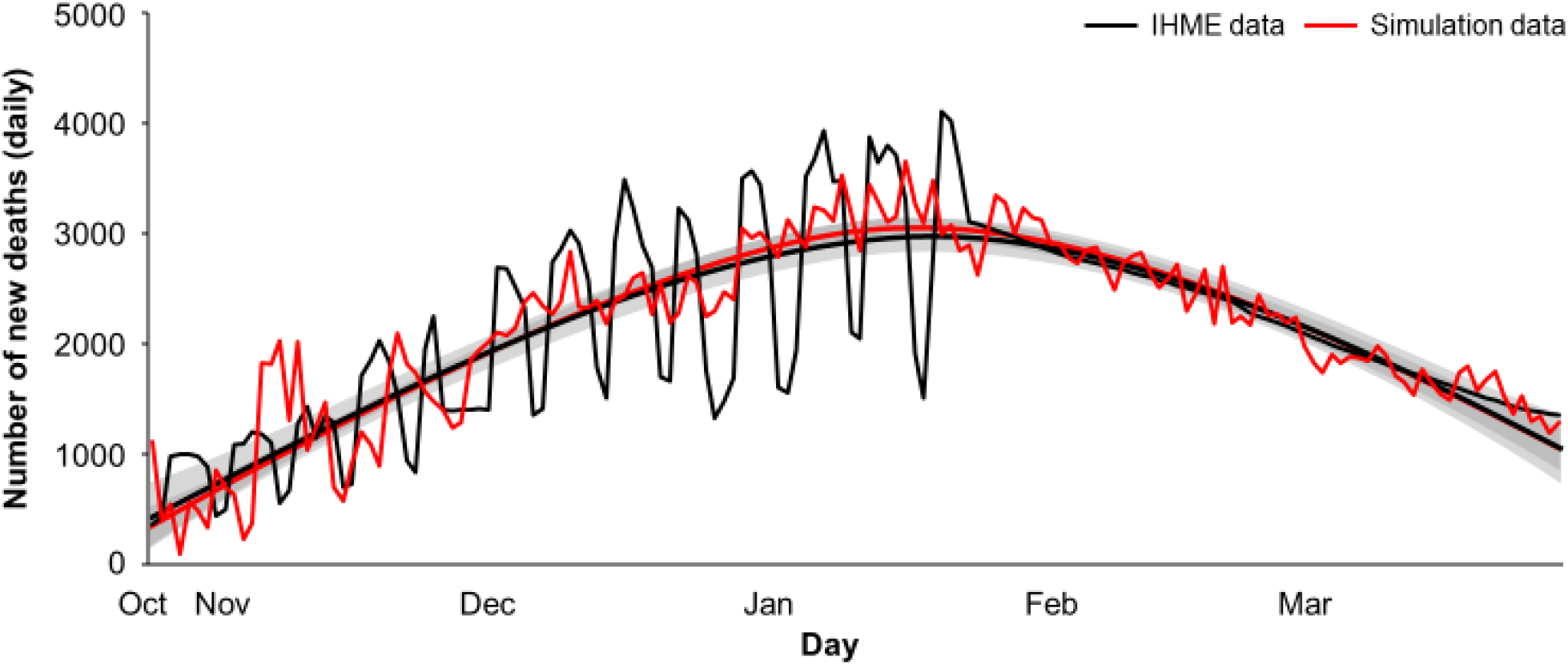
Calibration of the agent-based model using real-world mortality data. The model was calibrated to capture the daily distribution of deaths over a time period that reflected a more aggressive pandemic phase, from October 26, 2020 to April 4, 2021. When the agent-based model was fit to the observed distribution of deaths, the model tracked to the observed mortality curve. Observed data are from the Institute for Health Metrics and Evaluation (IHME).

### Evaluating impact of interventions on infections and mortality

Using our model assumptions (appendix p 24), we conducted simulations with the ABM to determine the effects of various mitigation strategies in combination under different scenarios during a more aggressive phase of the pandemic (October 26, 2020 to April 4, 2021). Multiple NPIs (e.g., travel bans, school and workplace closure, restriction of visitors to nursing homes, social distancing, facemasks, staying at home, and contact tracing) have been implemented in the US to mitigate the pandemic. In the current analysis, the aggregate impact of NPIs on cumulative infections and deaths in the absence of any pharmaceutical intervention comprised the base case. All subsequent simulations evaluating vaccine and mAbs were conducted on a background of NPIs.

All simulations with vaccines, including scenarios with mAbs, assumed a vaccine dosing interval of 25 days with efficacy of 52% and 95% after the first and second doses, respectively^18^ ; vaccine protection was assumed to start 7 days after the first dose. Since vaccine doses are limited over the simulation period, we prioritized agents who are either ≥65 years of age, living in nursing homes, or are medical workers targeted for vaccination; additional doses are distributed to agents ≥60 years of age or critical workers with greater social mixing, and therefore at higher risk of exposure.

Viral load served as proxy for infectiousness,^19^ and SARS-CoV-2 infectiousness was assumed to be proportional to the decimal logarithm of viral load in excess of 100 copies/mL, beginning two days following the first two days post-infection (latent period).^17,20^ A similar approach has been reported for influenza.^21,22^ To capture population variability in viral load profiles with and without mAbs, a viral kinetic model was used (appendix p 5),^23^ with the assumption that viral loads were highest at symptom onset, consistent with studies indicating that pre-symptomatic individuals are responsible for a large proportion of virus transmission.^11,24^ In our model, the median duration of infectivity in the absence of treatment was 9 days (range of 2-18 days). The impact of mAbs on median duration of infectivity varies with time of treatment; the reduction is 88% if administered 1 day after infection, 38% if treated at day 5 (time to symptom development), and 0% at day 10 (appendix p 26). For symptomatic patients we assumed 70% reduction in hospitalization and mortality risk.^7^

Since the current supply of mAbs is limited, the simulations assumed full utilization of 300,000 doses per month starting in January 2021, for a total of 900,000 doses, which reflects current manufacturing capacity from a single manufacturer. We also conducted a sensitivity analysis to interrogate the impact of a larger supply (600,000 per month for a total of 1.8 million doses), reflective of the expected supply from all manufacturers.

Assuming a 15% vaccine rollout, simulations were conducted with mAbs as both active treatment and PEP in a ratio of 1:2 to reflect PEP dosing at half of active treatment; PEP was defined as empiric administration to an unvaccinated person (agent) exposed to the virus through close contact but who does not present with symptoms and is of unknown COVID-19 status. Use of mAbs as PEP also assumed availability in a more convenient subcutaneous formulation for administration at a dose half that used as active treatment.^5^ Since the proportion of the US population vaccinated will increase over time, sensitivity analysis were conducted to determine the impact of increasing vaccine rollout from 15% to 30% and 47%, while keeping mAb supply constant at 300,000 and 600,000 doses per month, regardless of the increase in vaccinations.

Scenarios were also simulated incorporating rapid diagnostic testing for use in the home setting, such as the one from Ellume, which has been approved for emergency use in the US.^25^ The assumption was that patients would test themselves within 3 days (±1 day) of exposure, and if positive for COVID-19, would initiate early treatment that same day. With such testing, PEP may be more appropriately considered an early TasP strategy.

## Results

In the base case, there were approximately 103 million cumulative infections and 338,000 cumulative deaths over the simulation time period. Vaccine rollout of 15% averted almost 6 million infections and 43,000 deaths (figure 3, blue bar), corresponding to reductions of approximately 6% and 13% in transmission and mortality, respectively. In an analysis that increased the dosing interval from 25 to 60 and 90 days, these effects were relatively constant (appendix p 29), suggesting that real world deviations from the assumed dosing interval are unlikely to affect model predictions.

**Figure 3.**
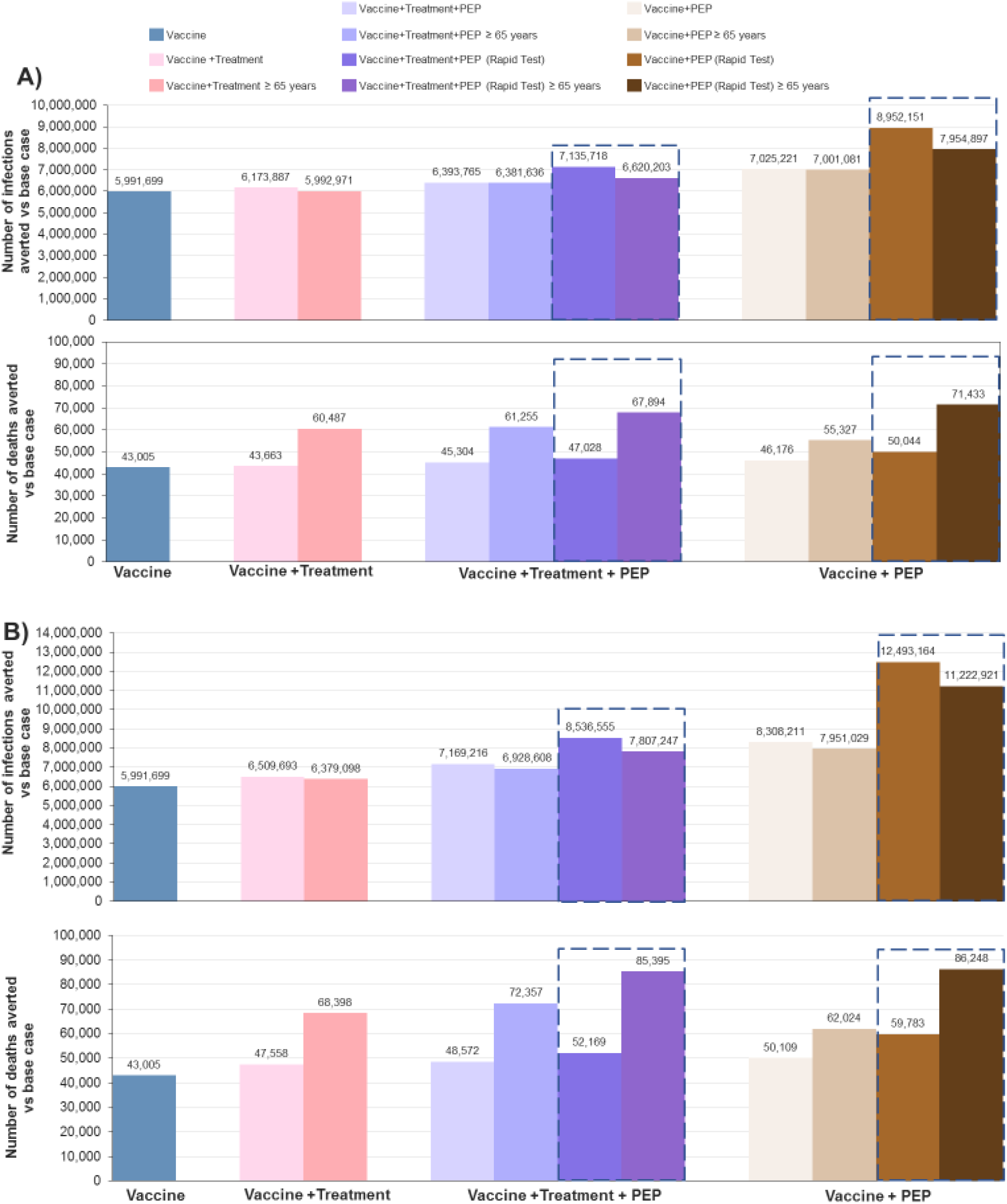
Simulations of the impact of monoclonal antibody treatment and prophylaxis in combination with vaccines (15% rollout). Simulations were conducted using the agent-based model to determine the contributions of different mitigation strategies on disease transmission (cumulative infections and deaths) during the aggressive phase of the pandemic (October 26 2020 to April 4, 2021). Monoclonal antibody supply from January 2021 was 300,000 doses/month (A) and 600,000 doses/month (B; sensitivity analysis), with total supply of 900,000 and 1.8 million doses, respectively. Results are presented as the number of infections or deaths averted relative to a base case of an aggregate of non-pharmaceutical interventions, which was characterized by 102,946,388 cumulative infections and 338,222 cumulative deaths over the time period. The colored columns reflect distinct paradigms, with shading indicating different scenarios within the paradigm. The columns enclosed by broken lines additionally incorporate the use of rapid diagnostic tests.

Model simulations with mAbs as active treatment (figure 3A, pink bars) in combination with vaccines show overall effects that are similar to vaccine alone. However, specifically targeting active treatment to those ≥65 years of age results in approximately 40% more averted deaths than with vaccine.

mAbs as both active treatment and PEP (figure 3A, purple bars) provide an incremental benefit of 7% relative to vaccine in the cumulative number of averted infections. However, targeting individuals ≥65 years of age resulted in a 41% increase in the number of averted deaths relative to vaccine alone.

Shifting allocation of mAbs solely for use as PEP further increased their benefits (figure 3A, brown bars); the number of averted infections (7 million) was higher by approximately 17% relative to vaccine (6 million), while at the same time reducing mortality by 7%. When an older population is specifically targeted, a 29% reduction in mortality (55,000 averted deaths) is achieved relative to vaccine (43,000 averted deaths), although additional benefits did not extend to infections averted.

When mAbs were used as both treatment (i.e., symptomatic infections) and PEP under conditions of rapid testing, the number of averted infections was 20% higher than vaccine alone, and when mAb allocation was shifted exclusively to PEP the number of averted infections was 50% higher (figure 3A). Targeting those ≥65 years of age provided smaller benefits in reducing transmission, but the number of averted deaths increased by 58%-66% (figure 3A).

Doubling the supply of mAbs from 300,000 to 600,000 per month (figure 3B) with full utilization resulted in benefits relative to vaccine that were greater than observed at the lower drug supply (figure 3A). Using rapid testing and allocating mAb to PEP at the higher supply averted approximately twice the number of infections (12 million) than with vaccine alone, with a 45% reduction in mortality (figure 3B). Similarly, targeting the elderly averted more than twice as many deaths and 89% more infections than vaccine (figure 3B).

Vaccine effects on reducing infections were proportional to the extent of vaccine rollout within the ranges we considered (Figures 3, 4; appendix p 18); infections averted increased from approximately 6 million to 12 and 18 million as vaccinations increased from 15% to 30% and 47%, respectively. mAbs consistently avert substantially more infections and deaths than vaccine alone regardless of the proportion of the population vaccinated (figures 3, 4; appendix p 18). The same pattern was observed in all scenarios: higher numbers of averted infections and deaths when mAb allocation is shifted to PEP, amplified effects when combined with rapid testing, and a sensitivity to mAb utilization. For example, even at 47% vaccine rollout (appendix p 18), monthly mAb supplies of 300,000 and 600,000 used as early TasP (i.e., PEP combined with rapid testing) averted 1·95 million and 3·82 million more infections, respectively, than vaccine alone. Similarly, using this scenario and targeting an older population results in approximately 17,000 and 25,000 more averted deaths at the two mAb supply levels, respectively.

**Figure 4.**
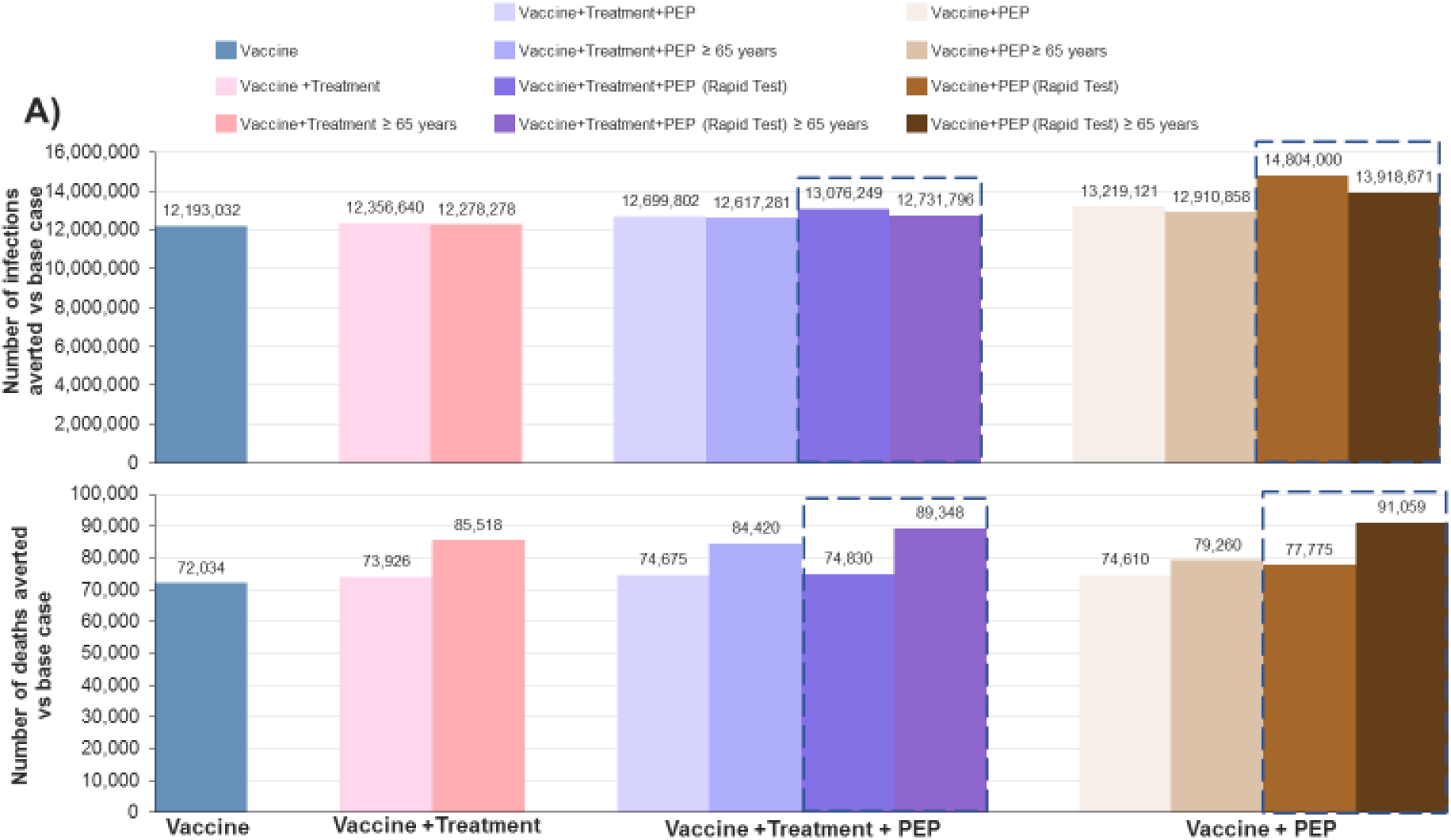
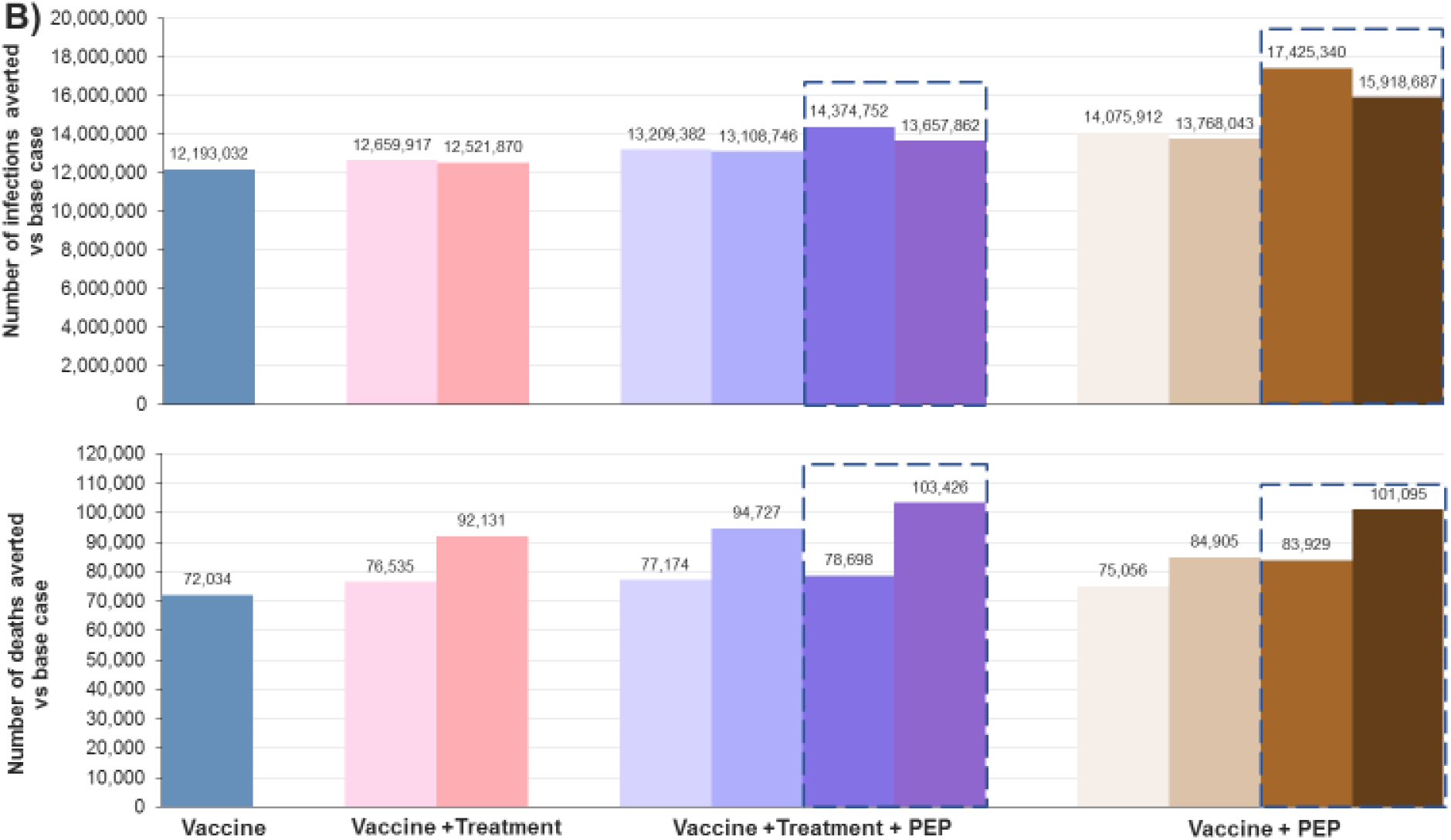
Sensitivity analysis of the impact of monoclonal antibody treatment and prophylaxis in combination with a 30% vaccine rollout. Simulations with the agent-based model were conducted under the same conditions as the main analysis but assuming a 30% vaccine rollout that was prioritized to those who are ≥65 years of age, living in nursing homes, or are medical workers, with additional doses distributed to those ≥60 years of age or essential workers with greater social mixing. Results are presented as the number of infections or deaths averted relative to a base case of an aggregate of non-pharmaceutical interventions (102,946,388 cumulative infections and 338,222 cumulative deaths) based on monoclonal antibody supply from January 2021 of 300,000 doses/month (A) and 600,000 doses/month (B). The colored columns reflect distinct paradigms, with shading indicating different scenarios within the paradigm. The columns enclosed by broken lines additionally incorporate the use of rapid diagnostic tests.

## Discussion

Modeling-derived insights are increasingly important from the perspectives of public health and policy decision-making given the rapid course of the pandemic and advances in treatment and vaccination. We therefore implemented a modeling and simulation approach to estimate the impact of mAbs in combination with other interventions on COVID-19 infections and mortality. While the use of mAbs as treatment has generally focused on reducing the risk of COVID-19 disease progression at the level of the individual patient, results of our model indicate an expanded role for mAbs including in combination with vaccines. This role encompasses their empirical use as PEP for reducing overall transmission and deaths, and when combined with rapid testing suggests an effective early TasP intervention.

Our base case was the aggregate impact of NPIs. While NPIs can blunt transmission,^2,3^ they require strict and prolonged adherence that may be difficult to implement nationally due to potential social and economic consequences, suggesting the importance of additional strategies to reduce transmission even in the presence of vaccines. Widespread vaccination is essential for achieving the long-term goal of herd immunity, and our results confirm a vaccine program provides quantitative benefits on a background of NPIs by reducing the number of infections and deaths. Other modelling studies have provided evidence of reduced transmission with this paradigm and have also emphasized the importance of adherence to NPIs even when implementing a vaccine program.^26,27^

Our model additionally provided evidence that reducing infections with use of mAbs, and by extension potential use of other pharmaceutical interventions, is critically dependent on logistics including time to treatment initiation. Rapid diagnostic testing to accelerate identification of infected agents for early mAb administration further amplified the benefits of mAbs. Pairing such testing with a readily accessible PEP regimen is expected to offer a substantial impact on both disease transmission and clinical outcomes by identifying pre-symptomatic individuals who are major contributors to transmission,^11-14^ thus facilitating mAb administration earlier than when used as active treatment. Earlier treatment consistently resulted in greater reductions in transmission and mortality as also indicated by the increasing numbers of averted infections and deaths as mAb allocation shifted from active treatment alone, to treatment + PEP, to use only as PEP.

When targeted to those ≥65 years of age, the benefits of mAbs were greater on mortality than on transmission, with similar patterns observed at all levels of vaccine rollout. This differential effect is not unexpected, since older patients are at higher risk of death, but less likely to transmit the virus. However, since this population is prioritized for vaccination, the incremental difference in averted deaths relative to vaccine was diminished as vaccine rollout increased. More recent projections of mortality from the Institute for Health Metrics and Evaluation (IHME)/University of Washington at Seattle indicate that while the number of deaths over the next several months will continue to decrease, 58,368 deaths are still expected to occur between April 13 and August 1 2021.^28^ A supplemental model simulation calibrated to these latest IHME mortality projections suggest that deployment of 1.25 million doses of REGEN-COV with prioritization to those ≥ 65 years old could avert 24,650 of the deaths (approximately 42%) during this time period, assuming deployment of 75% of the total drug supply during the first 50 days to temporally coincide with the peak of mortality.

Benefits of mAbs for reducing the pandemic burden were robust at all levels of vaccine rollout, especially when used as early TasP or specifically targeted to an older population, who may also serve as a proxy for other high-risk groups, i.e., younger individuals with multiple risk-related comorbidities. The observed relationship between vaccine rollout and averted infections is consistent with the reported association between vaccination and reduced transmission in a community setting.^29^ Regardless of the proportion vaccinated, mAbs substantially reduced infections and mortality relative to vaccine, although the magnitude of the difference was attenuated as the proportion of population vaccinated increased. These results emphasize the utility of mAbs even as herd immunity is approached; as the effective reproductive number, R(t), approaches 1, modest incremental benefits may still provide momentum in further reducing infections, thereby bringing R(t) below 1.

The estimated impact is also sensitive to mAb supply, with additional supply leading to larger benefits. There are two important implications regarding this observation. First, reducing logistical barriers to access and use will amplify the reduction in transmission and improve patient outcomes. Currently, mAbs are not approved for prophylaxis and are underutilized for treatment. This underutilization may arise from several sources, including lack of clarity of their potential contribution to reducing transmission. Our model suggests that broader use of mAbs for prophylaxis and early TasP can specifically contribute to reducing the pandemic burden when included as part of current mitigation policies. Second, the simulations can provide guidance on allocation of resources, and potentially lay a groundwork that can be applied to determine strategies for future pandemic preparedness. While mAbs provide an example of how an anti-viral therapy can be used as a mitigation strategy, the model is applicable to any intervention that can be used adjunctively with vaccination.

Study limitations include that these simulations reflect a specific time period of the US epidemic and that seasonal effects were not specifically considered. Another limitation is our assumption around vaccine effectiveness after the first dose, despite some recent studies suggesting that it may be higher.^30,31^ While our assumption was based on data available at the time the model was developed, the sensitivity analysis on increased vaccine rollout provides additional evidence of both vaccine efficacy and the incremental benefits that may still be achieved with mAbs. In addition, our model assumed uniform mAb distribution throughout the simulation period, and we did not consider the potential for added benefit using a surge distribution paradigm temporally coinciding with a phase in which the rate of mortality is increasing (e.g., late October 2020 through January 2021; figure 2). We also did not consider potential VOC when simulating the impact of mitigation strategies on the pandemic. As mutated viruses are indicative of a form of rapid, multistage evolutionary jumps (saltational evolution), this is an important component that may need to be characterized in future model iterations as data become available. The modular nature of the ABM makes it amenable to the potential occurrence of saltational evolution in this and future pandemics, for example by inclusion of a “viral resistance module”.

In conclusion, we present the results of an ABM, demonstrating how anti-viral mAbs may be used in combination with vaccines to suppress SARS CoV-2 virus transmission and improve clinical outcomes. These findings can be extrapolated to other anti-viral interventions with similar efficacy profiles to anti-spike mAbs. While vaccines may ultimately promote widespread herd immunity, the results of our model suggest that the near-term use of mAbs for early treatment and PEP, in combination with vaccines, provides additional public health benefits by reducing SARS CoV-2 transmission and related mortality, relative to vaccines. Increasing drug supply and utilization while lowering logistical barriers to access are integral to such mitigation strategies in addition to targeting specific populations at higher risk (e.g., ≥65 years of age) and increasing use of rapid testing. The benefits of mAbs appeared to be robust even as vaccine rollout increased, and these benefits are likely to be enhanced in scenarios where rapid uptake of vaccine utilization is not feasible. These results may help guide resource allocation and health policy decisions for COVID-19 management, and may additionally be applicable to future pandemic preparedness.

## Supporting information

Author COI

Author COI

Author COI

Author COI

Author COI

Author COI

Author COI

Author COI

Author COI

Author COI

Author COI

Author COI

Author COI

Author COI

Author COI

Author COI

## Data Availability

All data are available in the main text or the supplementary materials.

## Contributors

MAK, AK, and PFS conceived the study. LQ, MAK, AK, PFS, WW, KP developed and implemented the model. All authors participated in generating and interpreting the data. All authors contributed to the writing, review, and editing of the manuscript, and approved the work for publication.

## Data sharing

All data are available in the main text or the supplementary materials.

## Acknowledgments

This study was funded by Regeneron Pharmaceuticals, Inc. Writing support during manuscript development was provided by E. Jay Bienen, PhD, and was funded by Regeneron Pharmaceuticals. The authors thank Prime, Knutsford, United Kingdom, for manuscript formatting, which was funded by Regeneron Pharmaceuticals, Inc.

