## Supplemental Materials for "Monoclonal Antibody Treatment, Prophylaxis and Vaccines Combined to Reduce SARS CoV-2 Spread"

### Materials and methods

### Description of the agent-based model

The simulations performed in this study utilized an agent-based computational model (ABM). The ABM allowed us to simulate the spread of SARS-CoV-2 in the US, and evaluate pharmaceutical intervention strategies, while accounting for heterogeneity among agents. Details regarding model implementation are described in this section.

#### Population generation and contact structure module

The individual (agent)-level data were derived from American Community Survey census statistics. We used the latest available 5-year aggregated census results (2018), or the most recent data in cases where these were not available (input tables detailed in Table S1).

We first derived countywide populations, stratified by age and sex, that resembled the US and then generated the following characteristics using empirical probability distributions provided by census data:

1) Residency into grouped quarters (student housing, nursing home)
2) Employment status and types of occupations
3) School attendance

An implicit assumption of our population generation process was that the age- and sex-stratified estimates sufficiently captured correlations among the characteristics of interest. After collecting individual-level characteristics, individuals not living in group quarters were grouped into households (Table S2) following the household structure from census data. Finally, individuals were allocated to the 3,143 counties and county-equivalents of the US based on US Census Table B01001, in terms of counties of residence and work, and according to the reported county-to-county commuting workflows.

The generation process included (Fig. S1):

1) Table B01001 (all table numbers are those listed in Table S1) was used to generate a population whose size was equal to the full US population, with age based on the multinomial distribution available from the input table (23 age groups). Within each age group, actual age was assumed to follow a uniform distribution. Sex was age-stratified and drawn from a binomial distribution using reported data from the same table.
2) Table B14004 was used to identify individuals who were enrolled in higher education programs (college). This table stratified individuals into four age categories (15–17, 18–24, 25–34, and >35 years of age), by sex.
3) The National School Enrollment data were used to allocate type of schools for all individuals. Of note, these data were only available at the state level. Probabilities were first rescaled to account for discarding of individuals already enrolled in college. Agents could be enrolled in kindergarten, elementary school or high school. All sampling was performed using empirical multinomial distributions specific to age and sex.
4) Table B14005 and B14004 were used to refine state-level school sampling from step 3) to county-level in the subset of individuals aged 16–19 years old. First, the probability of being enrolled in school was drawn using a binomial distribution based on Table B14005, then the type of school was determined using multinomial sampling, based on Table B14004.
5) Table B14005 was used to derive probabilities of being part of the labor force for individuals enrolled in school and aged 16–19 years old. For each combination of sex, age group and school enrollment status, a temporary working status was drawn from a binomial distribution (employed or not employed).

6) Table B23001 was used to determine age- and-sex-specific probabilities of being allocated into different work categories. First, probabilities were normalized to one by discarding ages below 20 years old, to account for county-level work assignments sampled at step 5. Second, multinomial sampling was used to categorize agents into the following work groups: "Armed Forces", "Employed Civilian", "Unemployed Civilian" and "Not Part of Labor Force". The latter two categories were subsequently regrouped.
7) Table B23001 was used for agents aged 16–19 years old, marked as "working" at step 5, to draw work categories as described at step 6. Multinomial sampling from empirical data was used.
8) Table B08006 the source for determining if workers were home-based, using binomial sampling.
9) Table B24010 was used to refine work types. Individuals were categorized as working "Client" jobs, "Office" jobs (including the "Armed Forces") or "Medical" jobs.

The following procedure was used to generate a social network, i.e., clusters of individuals interacting closely with each other.

1) Based on a scaling factor, individuals were sampled from the previously generated population. The scaling factor was used to achieve a sample of specified size, making the epidemic simulation computationally tractable while preserving representativeness of the agents' characteristics of interest. Population diagnostics comparing the distribution of these characteristics were performed at the state and national level to assess validity of the synthetic population. The national-level diagnostics are provided in Fig. S2 through Fig. S5.
2) County-level communities were generated as subsets of ~2000 agents.[1] After the expected number of communities were obtained, uniform sampling was used to allocate community IDs to each individual. Within each community, 4 distinct neighborhoods were created, using uniform sampling, each of target size ~500.[1]
3) Tables P42 and B26101 were used to randomly allocate residency to student housing for individuals enrolled in colleges. The target size of the student housing was 280, estimated from the housing data of Columbia University (https://housing.columbia.edu/, accessed on 20 July 2020). After estimating student housing, multinomial sampling was used to allocate individuals into these student housing groups.
4) Table B26101 was used to identify individuals residing in nursing homes in a process similar to step 3 for student housing. The target size for nursing homes was 100.[2]
5) Table B11016 provided county-level estimates of household size distributions and was used to allocate agents to households. In this step, we also used Tables B09021 (proportion of young adults living with their parents) and B23008 (distribution of number of children living with a single adult).
6) Schools were processed similarly with regard to how individuals were allocated into group quarters at steps 3 and 4. The target sizes of kindergarten, elementary, middle school, high school and college were 14, 79, 128, 155 and 155, respectively.[1,3]

In the above, the actual size of the community groups varied around the target. For each type of group $c_k$, a number $N_{c_k}$ of groups was calculated so that, on average, no group could have less than 30% of the target size $N^*_{a,c_k}$, with $N_{a,c_k}$ representing the number of agents belonging to group $c_k$: Equation (1) details the calculation of $c_k$. After $N_{a,c_k}$ was calculated, agents were randomly assigned into each group with uniform sampling.

$$N_{c_k} = \begin{cases} \left\lceil \frac{N_{a,c_k}}{N^*_{a,c_k}} \right\rceil if\ N_{a,c_k}\ mod\ N^*_{a,c_k} \geq 30\% * N^*_{a,c_k} \\ \max\left(1, \left(\left\lceil \frac{N_{a,c_k}}{N^*_{a,c_k}} \right\rceil - 1\right)\right) otherwise \end{cases} \quad (1)$$

After inputting the population data into the C++ stochastic simulator, each individual had the following baseline characteristics: ID, age, sex, household, neighborhood, community, employment status, school status, workplace and school. Each individual also had one health status with respect to COVID-19. This status was updated at each simulation time step (1 day).

As outlined above, individuals were associated through five contact contexts (i.e., household, workplace, school, neighborhood, and community), also called "mixing group", and could be infected by other individuals in the same mixing group. The household, school and workplace were specified transmission pathways, with household as the primary pathway. Communities and neighborhoods represented places where people could have casual contact with others outside of the household, school or work (unspecified transmission pathways, e.g., during shopping or social interactions). The individuals and their associations formulated our base social network (Fig. S6) where daily activities occurred and SARS–CoV–2 virus spread.

#### Movement module

In addition to the regular daily activities occurring inside the base social network, individuals could have infrequent and irregular long-distance, short-term domestic travel. The long-distance travel component was required because we aimed to simulate the spread of epidemic throughout the US. The datasets used in the movement module are listed in Table S3. Implementation of long-distance, short-term trips was similar to other models in the literature.[1,3] Each non-hospitalized individual had a daily and age-specific probability to start a new trip that could last between 1 and 12 days (Table S4 and Table S5). The trip destination was a randomly selected household inside the destination state and was dependent on the household's state; most trips occurred within the household's state. As an example, Fig. S7 presents the destination states for individuals in Alabama.

#### Virus transmission and disease progression module

The probability that a susceptible individual (S) was infected per day P(t) was dependent on: (1) age of the S; (2) risk from imported cases; and (3) all infectious individuals in the same mixing groups as S. We calculated this probability based on the concept of person-to-person transmission.[1,4] The formula for virus transmission is presented in Equation (2).

$$\mathrm{P(t)} = 1 - \left(1 - P_{imported}(t)\right) * \prod_i \left(1 - C_i * VL_i(t) * P_{trans}(t)\right) \quad (2)$$

$C_i * VL_i(t) * P_{trans}(t)$ represents the probability that one S individual was infected by one infectious, non-hospitalized individual *i* who belonged to the same mixing group. $C_i$ represented the probability of a sufficient contact for transmission during one time step (name as "contact probability"), dependent on the age and the mixing groups of S. $VL_i(t)$ represents the infectiousness of the infected agent, proportional to the decimal log of viral load in excess of 100 copies/mL. $P_{trans}$ (t) was a scalar used to adjust the overall transmission probabilities $C_i * VL_i(t)$ to obtain a longitudinal mortality curve for target simulation scenarios as discussed in the main article text. Note that in all simulation scenarios, $C_i * VL_i(t) * P_{trans}(t)$ did not exceed 1 (maximum value was 0.1747896). $P_{imported}(t)$ represents the probability that one S individual was infected due to the risk from imported cases (international passengers from outside the US). Since our analysis was not modeled on the initial phase of the pandemic, we focused on propagation of SARS–CoV–2 inside the US while assuming the risk from imported cases was negligible.

After acquiring an infection, COVID-19 progressed as shown in Fig. S8. After an exposed state (E), the infected individual had a probability to develop to a pre-asymptomatic state (Incu, the infectious stage of the incubation period) from which they might develop symptoms at a later disease stage. The time from infection to symptom onset was assumed to be 5 days,[5] including 2 days as exposed and not infectious,[6] and 3 days as pre-symptomatic and infectious. Two types of symptomatic infections (mild and severe) were modeled, and only severe state could lead to death (D) after passing the hospitalization state (Hosp). The parameters used in the disease progression module are described in Table S6 and Table S7.

#### Vaccination module

The vaccination module was implemented to mimic the US vaccination program to enable evaluation of the real-world effects of monoclonal antibody therapies. We modeled vaccines requiring two doses for full protection (the J&J single-dose vaccine was not authorized at the time the model was developed). Based on the recommendation of the Centers for Disease Control and Prevention (CDC), both susceptible (S) and recovered (R) agents could be vaccinated.

The model assumed different vaccine effectiveness before and after the second dose, with another parameter modulating the days between the two doses. We assumed a 25-day gap between the two doses, with real world effectiveness (assuming effectiveness as measured in randomized controlled trials) as 52% for days 7–25 and 95% after day 25 (second dose).[7] No assumption was imposed on the heterogeneity following a successful vaccination, thus individuals with successful vaccination were fully protected against all forms of the disease without loss of immunity (e.g., due to variants).

The vaccine supply varied by week as shown in Table S8, derived from longitudinal vaccine data from the CDC. Since the vaccine doses were limited and taking the CDC recommendations (https://www.cdc.gov/coronavirus/2019-ncov/vaccines/recommendations.html) into consideration, by default, we prioritized the vaccination to agents who were either ≥65 years old, those living in nursing homes, or medical workers, and additional doses were distributed to agents ≥60 years of age or working in client-facing jobs (Table S9).

#### Antiviral treatment module

Antiviral treatment with anti-spike monoclonal antibodies could reduce both the duration of viral shedding, and the probability of disease progression.[8]

The longitudinal viral load used to model the time-dependent infectiousness of each infectious individual ($VL_i(t)$ in Equation 2) was generated from a previously published target cell-limited model of SARS-CoV-2 viral infection dynamics[5] that used SARS-CoV-2 viral kinetics data pooled from 13 published studies, since the clinical trial data were not available when the ABM was developed. Following availability of clinical trial data,[8] the target cell-limited model[5] was calibrated to incorporate antiviral effects that best predict SARS-CoV-2 viral kinetic data with monoclonal antibody treatment based on data derived from the clinical trial of the Regeneron monoclonal antibody combination, REGEN-COV treatment.[8] The calibration process is described below.

Initially, the target cell-limited model was used to simulate antiviral effects by modulating various components of the viral life-cycle, including inhibition or enhancement of one parameter or a combination of viral kinetic parameters. The following antiviral effects were tested, and were evaluated by overlaying preliminary viral load versus time data following monoclonal antibody administration:

1) Inhibition of viral production rate by 70%, 90%, 95%, or 99%
2) Enhancement of clearance of infected cells by 1.5-, 2.5-, 4-, or 5-fold
3) Enhancement of free virus clearance by 1.5-, 2.5-, 4-, or 5-fold
4) Inhibition of viral production rate by 70%, 90%, or 95%, plus enhancement of infected cell clearance by 1.5-, 2.5-, 4-, or 5-fold

5) Inhibit viral production rate by 70%, 90%, or 95%, plus enhancement of free virus clearance by 1.5-, 2.5-, 4-, or 5-fold
6) Inhibition of viral production rate by 70%, 90%, or 95%, plus enhancement of infected cell clearance by 1.5-, 2.5-, 4-, or 5-fold plus enhancement of free virus clearance by 1.5-, 2.5-, 4-, or 5-fold.

Conducting these simulations determined that the combined effect of enhanced viral clearance of infected cells (by 1.5-fold) and reduced production (by 70%) provided predictions that broadly reflect the COVID-19 profile following monoclonal antibody administration.[8] We used the well-modulated target cell-limited model to simulate 100 viral load profiles (in the absence or presence of treatment, with treatment initiated at various days post infection) in order to capture between-individual variability in viral load. In the ABM, each infected individual was randomly assigned one of the 100 simulated viral load profiles.

The control profile was used when modeling the default infectiousness without the antiviral treatment. From the 100 simulated control profiles, we found that the average infectious duration was 9 days (SD = 5 days), ranging from 2 to 18 days and that the cumulative infectiousness (area under the curve [AUC]) in the first 5 days (from infection to symptom onset) accounted for, on average, 54% of the total value (ranging from 17% to 100%). The cumulative infectiousness during the pre-symptomatic stage (3–5 days post infection) accounted for, on average, 38% of the total value (ranging from 14% to 58%).

Once an individual received the antiviral treatment, the treatment profile was used to model their infectiousness. In the treatment profile, the viral load values that corresponded to the treatment administered from the third day post infection were directly used, while the viral load values of the treatment administered on the first 2 days post infection were generated from the control profile based on the assumptions of the treatment effects (Table S10). The reductions of the median infectious duration were calcuated from the simulated viral load profiles (Table S10). As an example, Fig. S9 shows one full viral load profile used in the ABM (including both the control and the treatment profiles) to illustrate reduction of infectiousness and its duration by treatment. If the drug was administered to any susceptible agent, we made a conservative assumption that the drug could prevent infection for a duration of 30 days.

Outpatient treatment could also reduce the probability of disease progression (Table S11), consistent with clinical trial data (under review at *The New England Journal of Medicine*). We assumed that the antiviral treatment only reduced the probability of progressing to the subsequent worse disease stage (Fig S8). For example, if an individual received the treatment with the health status as exposed (E), this individual would not progress to the incubation stage (Incu, also known as pre-symptomatic stage). Analogously, if an individual was treated at the severe stage, the probability of being hospitalized would be reduced by 70%.

### Estimation of the illness attack rate

The illness attack rate, which was needed for model calibration and simulation initialization, was estimated from the cumulative death data (Fig. S10), death distribution by age (Fig. S11), and the infection fatality ratio (Table S12). Detailed data sources are listed in Table S13. The total number of previously infected individuals, specific to age group, was simply calculated by dividing the cumulative deaths by the infection fatality ratio. For example, the illness attack rate for June 1, 2021 was calculated by dividing the cumulative deaths on June 23 by the infection fatality ratio. Table S14 presents the estimated illness attack rates for June 1, 2020 and November 1, 2020.

### Model calibration

In the calibration process, various model parameters were modulated using an adapted gradient descent method, i.e., assuming a linear relationship between the parameters and the outcomes.[1]

#### Calibration of the contact probabilities $C_i$

Calibration of the contact probabilities $C_i$ was location- (household, workplace, school, neighborhood, and community) and age-specific (0–18, 19–64, and ≥65 years), and was the first step of the calibration process. The calibrated parameters $C_i$ were assumed to represent the base contact structure in the population and were modulated by $P_{trans}(t)$ over time to represent the degree of aggressiveness of virus propagation. We calibrated $C_i$ using the illness attack rates for June 1 and November 1, 2020 (Table S14), since we considered that the behavior of individuals within the US had already adapted to the pandemic after strong non-pharmaceutical interventions (NPIs, e.g., shelter in-place and non-essential business closure) implemented between March and early June 2020, and that this time period reflected a mild and typical propagation of SARS–CoV–2 in the population.

Initial calibration of the contact probabilities adjusted the $C_i$ of the household to match secondary household attack rates by age group. Since the household was the primary transmission pathway in our model setting, the household was considered the primary source of infection. The other four pre-specified mixing groups (workplace, school, neighborhood and community) shared the remainder of infections. Table S15 presents the results of the calibration on the household secondary attack rate. Calibration results on the illness attack rates, shown in Table S16, are similar to those by age calculated from the cumulative death, death distribution by age and the infection fatality ratio by age (Table S14). The source of infection generated during the calibration process is shown in Table S17 and calibrated parameters $C_i$ are listed in Table S18.

#### Calibration of the disease progression parameters

After calibrating the contact parameters $C_i$, we varied the parameters of disease progression to match the infection fatality ratio (Table S12). As shown in Table S19, after the calibration, the overall and age-specific infection fatality ratios simulated from the model were similar to real-world estimates (Table S12). The calibrated parameters that were used in the disease progression module are listed in Table S6. We also noted from the simulation of baseline scenario (without vaccine and antiviral treatment), that the hospitalization rate of total infected individuals over the simulation period was approximately 2%; and the death rate among those hospitalized was approximately 18%, similar to observation data.[9,10]

#### Modulation of the contact probabilities over time

The last step of model calibration was to modulate the parameters $C_i$ by varying $P_{trans}(t)$ bi-weekly to match the longitudinal death observations as required by the target simulation scenarios. We calibrated ten $P_{trans}(t)$ to match longitudinal death data between November 16, 2020 and April 4, 2021 (using current projection data from the Institute for Health Metrics and Evaluation [IHME], Fig. S12). The model was initialized to match the death and the illness attack rate on October 26, 2020. The final ten $P_{trans}(t)$ values obtained from the calibration process are listed in Table S20. Detailed data sources are listed in Table S21.

### Antiviral treatment strategies

The active treatment, as a single dose, was provided to individuals with symptoms, on average 3 days post symptoms (SD = 1.4 day).

Post-exposure prophylaxis (PEP) was provided to individuals with at least one confirmed case in the household (close contacts). Individuals receiving PEP (0.5 dose) were either susceptible or on average 3 days (SD = 1 day) post-infection (without symptoms). The ratio of PEP used for non-infected to infected was approximately 3 (i.e., 25% of PEP was used on actually infected people; detailed results from simulations are shown in Table S22). When rapid test was available, the PEP was only administered to confirmed cases (100% of PEP was used on actually infected people) to describe an intervention that mimicked early Treatment-as-Prevention (TasP). Those patients were identified through contact tracing (share the same household, workplace, or school with a confirmed case). We assumed any test approved by FDA would have reasonable performance and did not make assumptions around test sensitivity or specificity.

#### Computing environment

All data management (including population generation) was performed using R (version 3.6.3, https://cran.r-project.org/bin/windows/base/old/3.6.3/). The stochastic simulator, developed in C++ under the Eclipse environment (Eclipse for Parallel Application Developers, https://www.eclipse.org/downloads/packages/release/juno/sr2/eclipse-parallel-application-developers), only loaded the well-prepared data and ran the simulations. OpenMP was used in the stochastic simulator to realize the parallel computing (multiprocessing programming) among simulation runs. The simulation scenarios were performed on an internal computing server of Certara (64-bit operation system, x64-based processor). There were 16 processors (Intel(R) Xeon(R) Gold 6132 CPU @ 2.6GHz 2.59 GHz) and 96.00 GB installed RAM. For each simulation scenario, we reported the average values of 50 realizations as final outputs. It took approximately 1 hour to process one simulation scenario of 50 realizations (duration of each realization from October 26, 2020 to April 4, 2021).

### Supplementary figures and tables

**Fig. S1** Flowchart of population and social network generation process

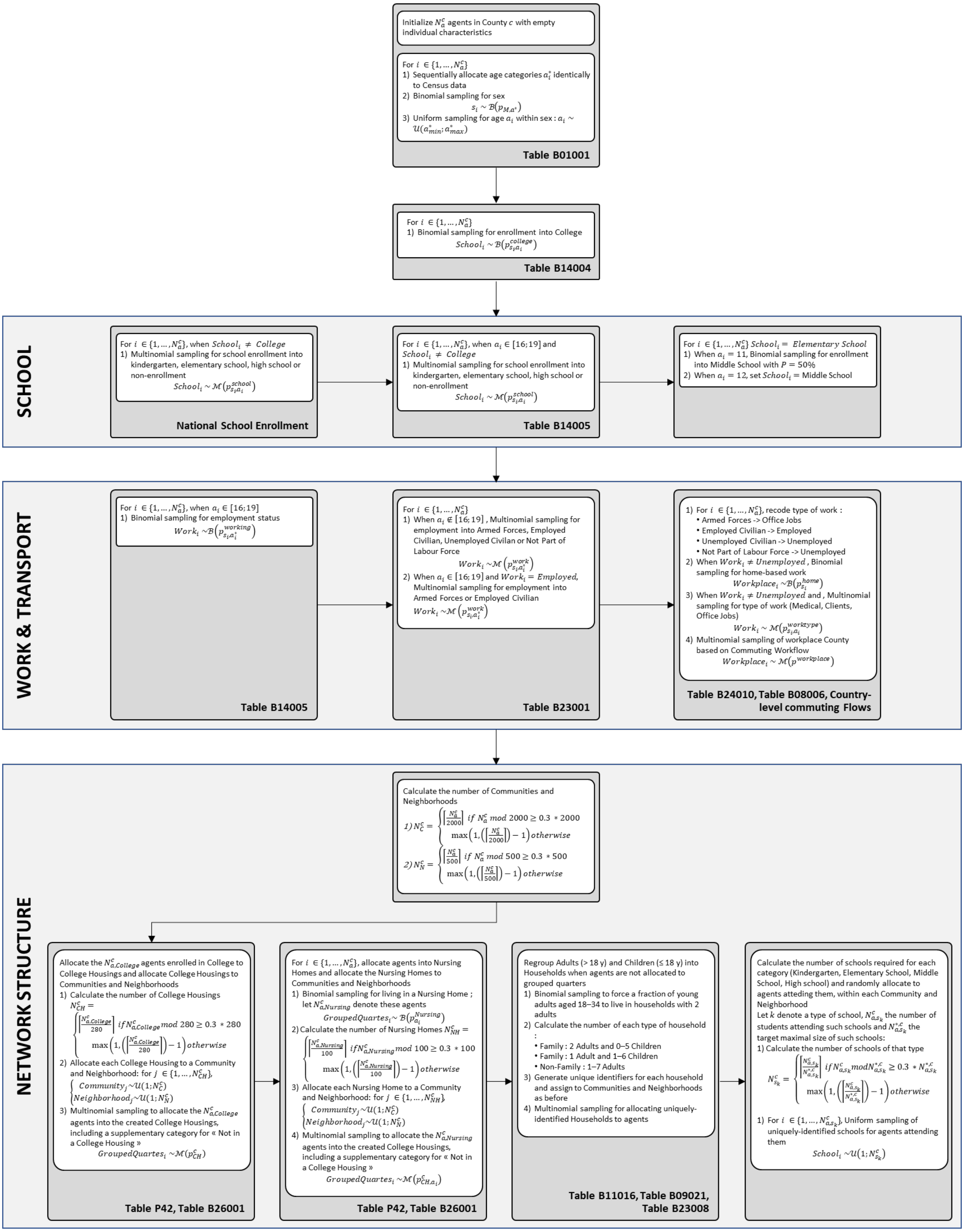

**Fig. S2** Population diagnostics graph for age-and-gender stratification.

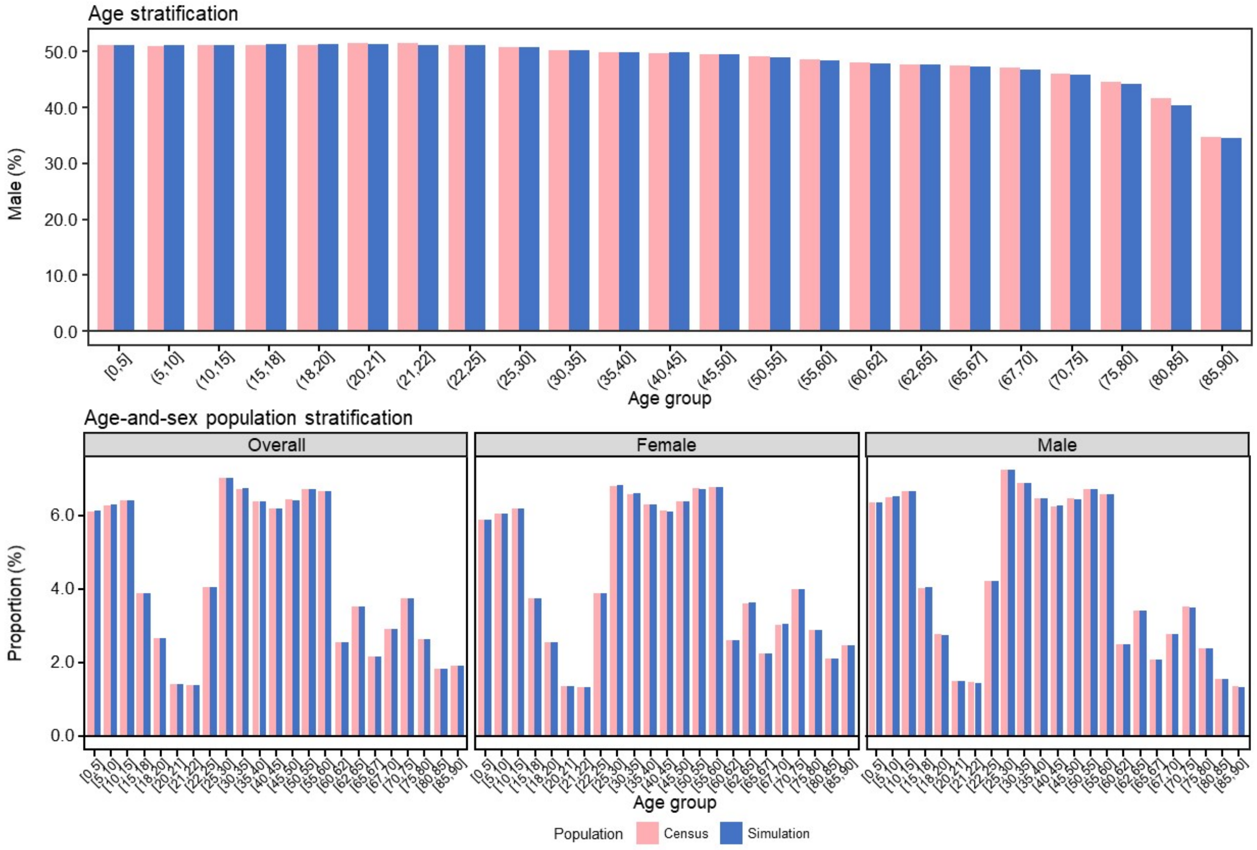

**Fig. S3** Population diagnostics graph for age-and-gender-stratified employment rate.

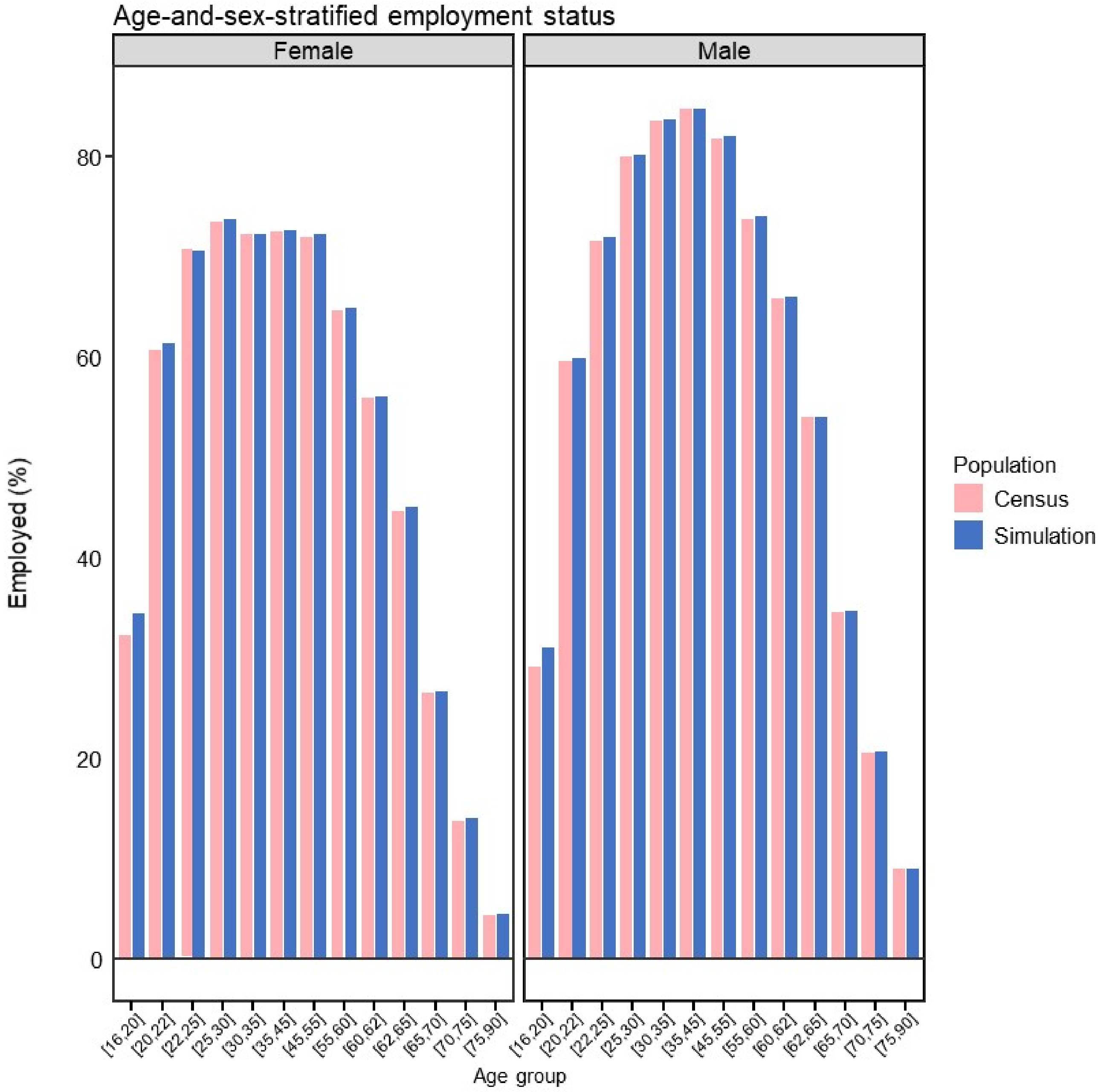

**Fig. S4** Population diagnostics for age-and-sex-stratified enrollment in schools.

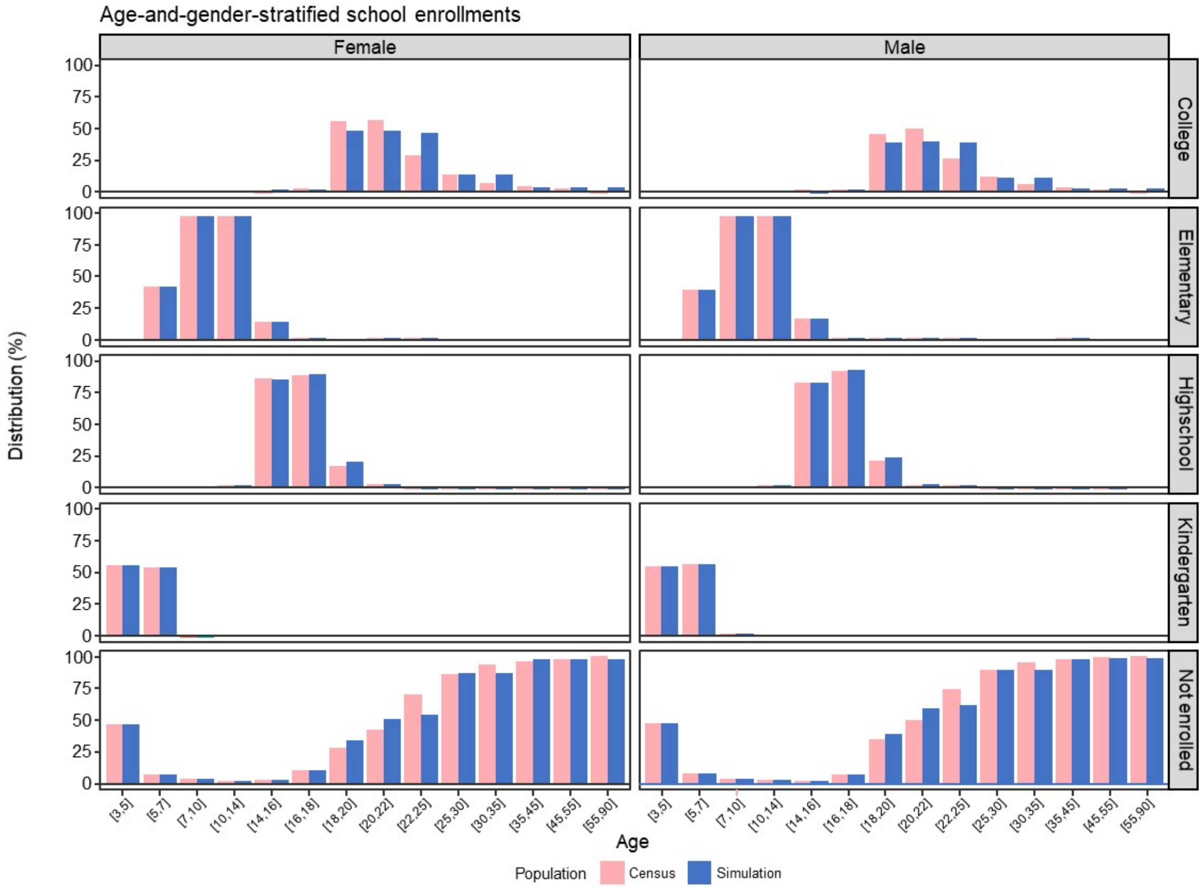

**Fig. S5** Population diagnostics for sex-stratified type of work in the employed population force.

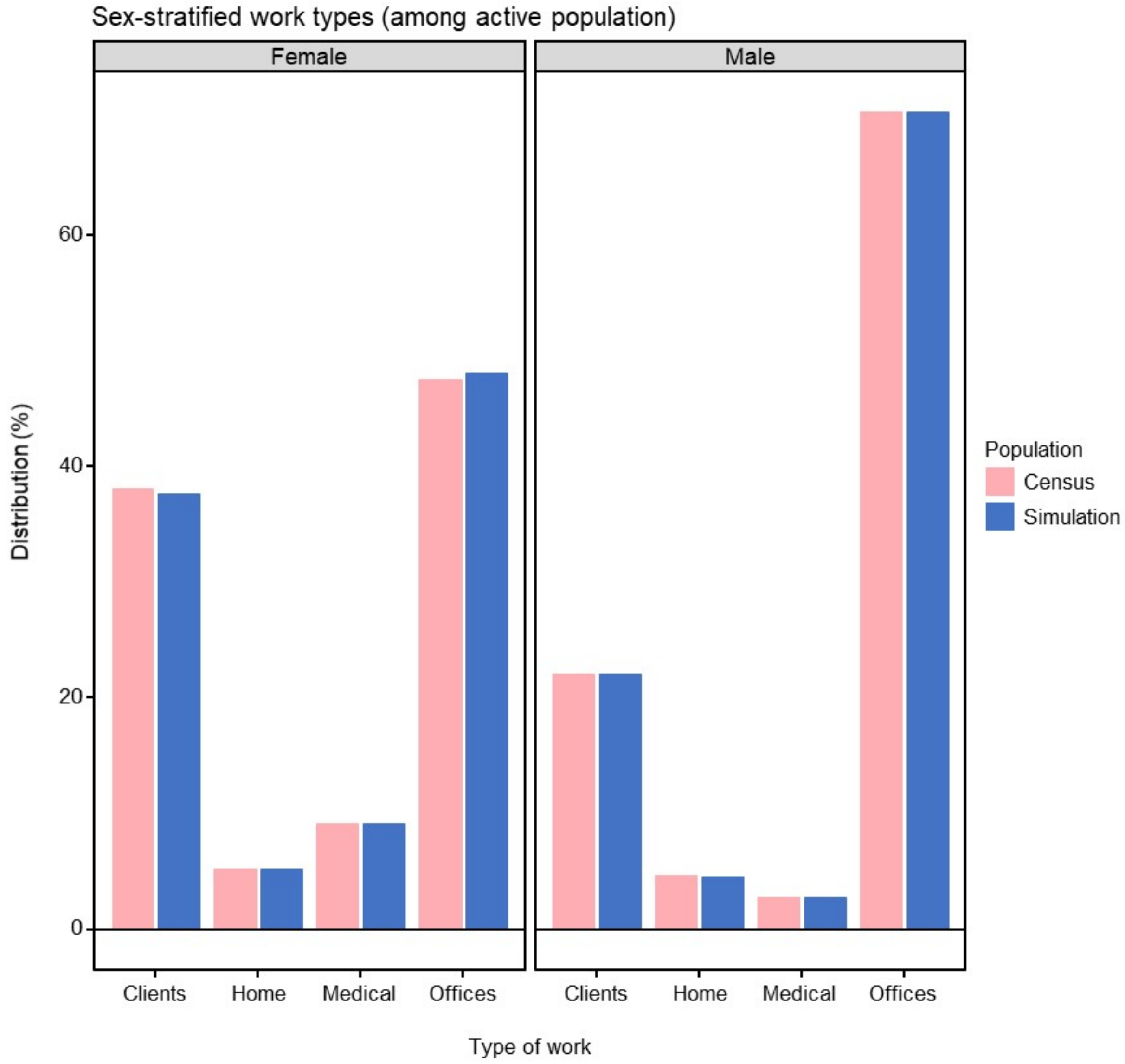

**Fig. S6** Base social network of the ABM.

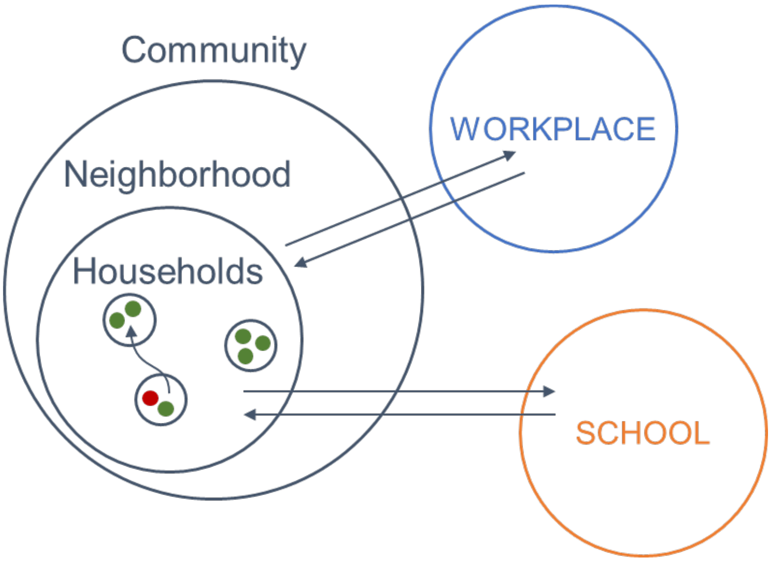

**Fig. S7** Top ten destinations of the long-distance short-term domestic trips of people from Alabama.

| Top destinations | Shares |
|---|---|
| Alabama | 41.4% |
| Georgia | 14.6% |
| Florida | 11.1% |
| Tennessee | 10.5% |
| Mississippi | 5.7% |
| Louisiana | 2.9% |
| North Carolina | 1.6% |
| Texas | 1.3% |
| South Carolina | 1.2% |
| Missouri | 1.1% |

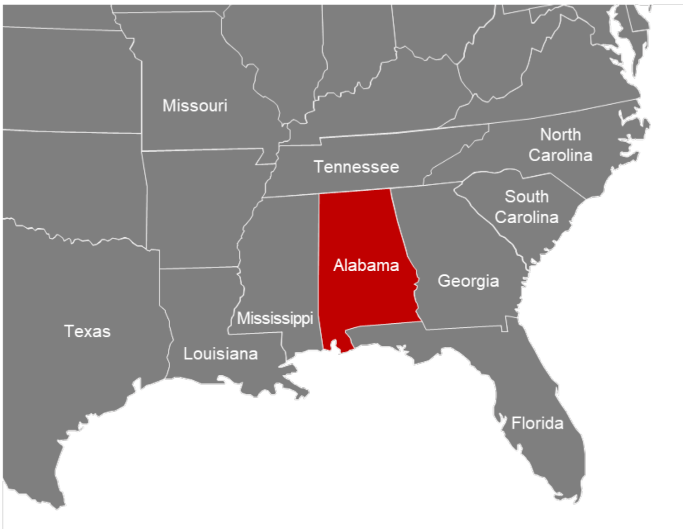

**Fig. S8** Disease progression after infection: (1) E: exposed and not infectious; (2) A: asymptomatic; (3) Incu: pre-symptomatic (the infectious stage of the incubation period); (4) Mild: symptomatic with mild symptoms; (5) Severe: symptomatic with severe symptoms; (6) Severe_rec: recovery period for patients with severe symptoms; (7) Hosp: hospitalization; (8) Hosp_rec: recovery period for hospitalized patients; (9) ICU: intensive care unit; (10) D: death; (11) R: recover from infection.

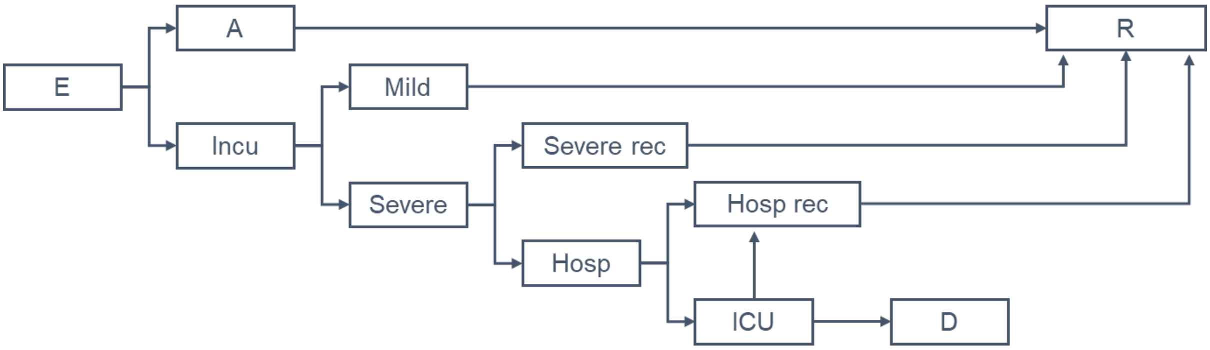

**Fig. S9** One of the 100 simulated viral load profiles. The control profile shows viral load values without antiviral treatment and other profiles show viral load values if treated on the infected day post-infection.

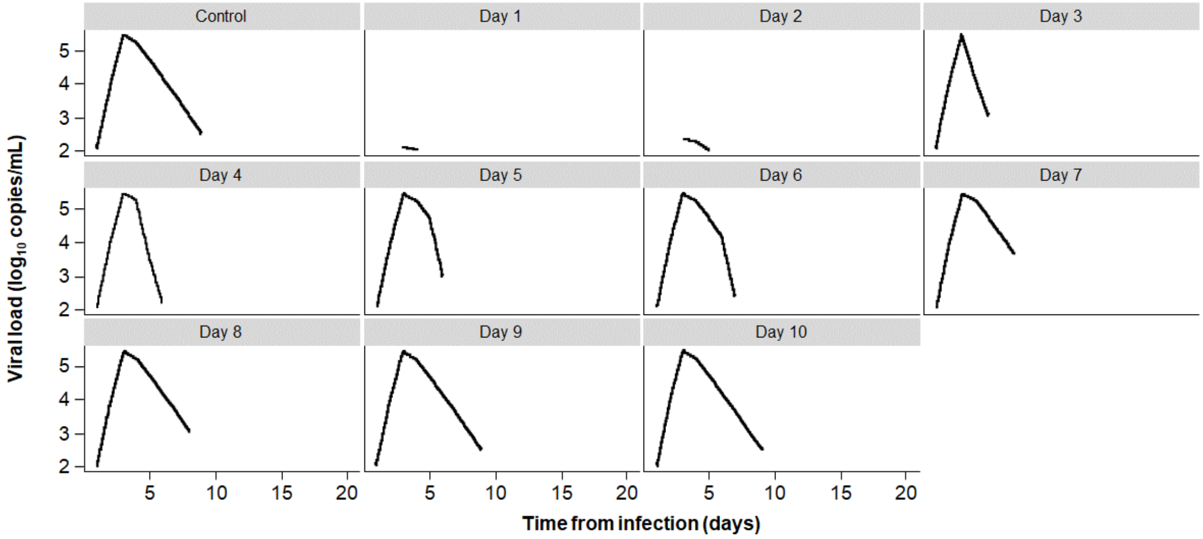

**Fig. S10** Cumulative death over time (CDC data, accessed on January 22, 2021, https://data.cdc.gov/Case-Surveillance/United-States-COVID-19-Cases-and-Deaths-by-State-o/9mfq-cb36).

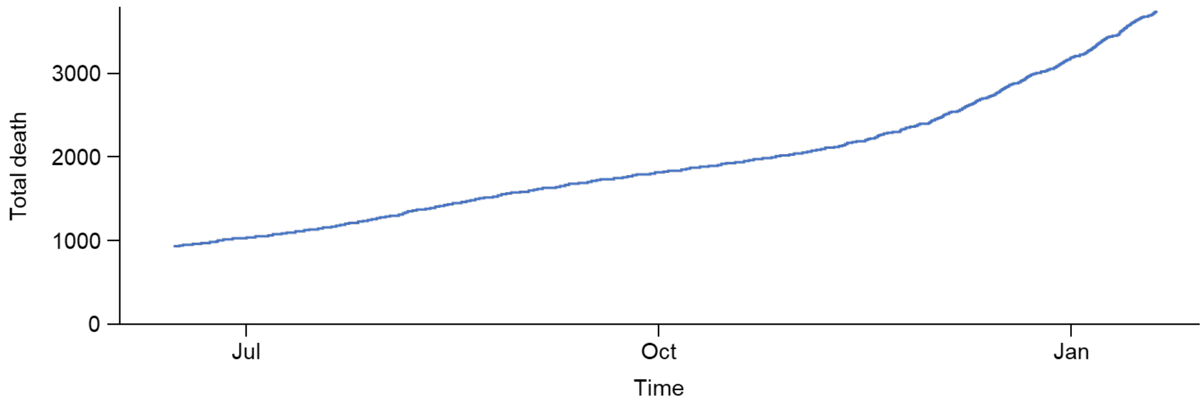

**Fig. S11** The death distribution by age group used in the simulations (from CDC on November 23, 2020, https://covid.cdc.gov/covid-data-tracker/#demographics).

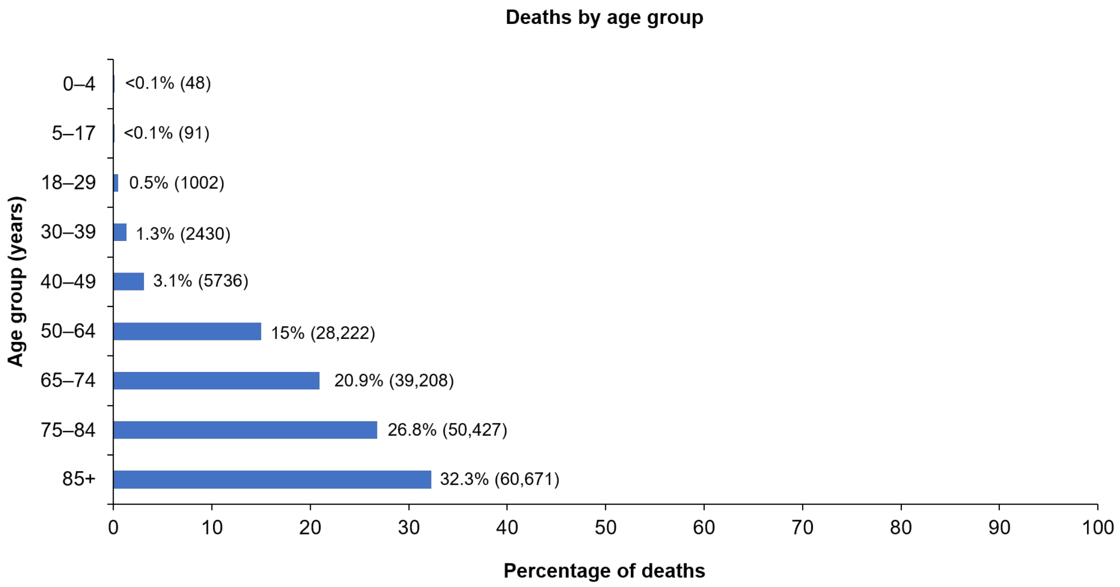

**Fig. S12** Longitudinal death observations (CDC data accessed January 22, 2021 and IHME data accessed January 28, 2021; data sources are listed in Table S21).

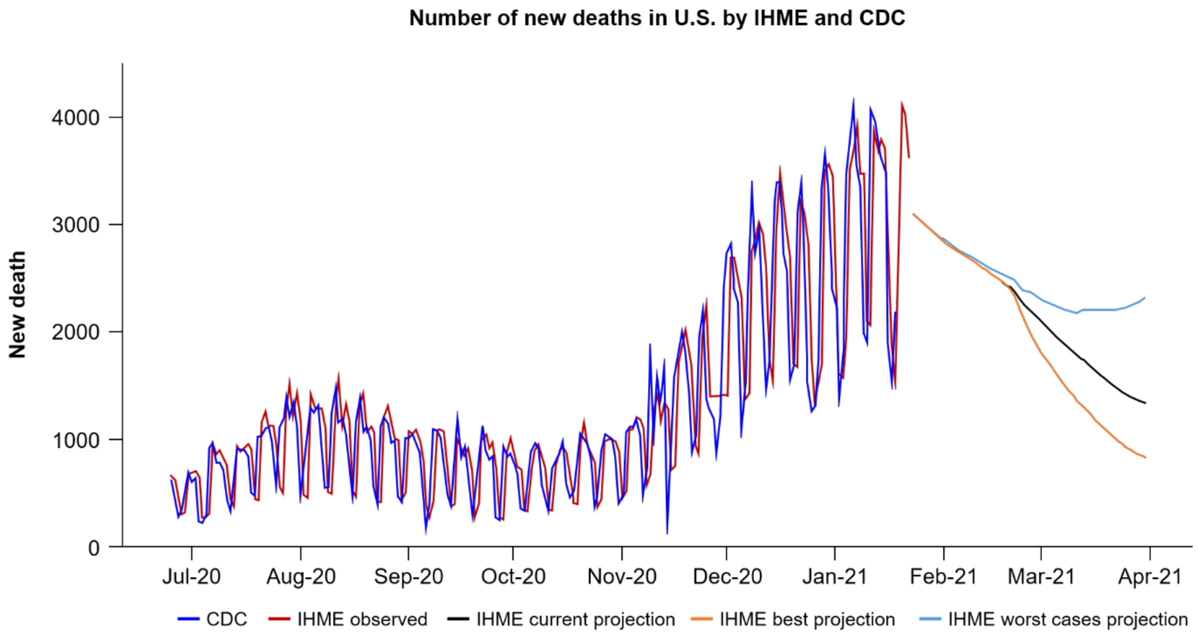

**Fig. S13** Sensitivity analysis of the impact of monoclonal antibody treatment and prophylaxis in combination with a 47% vaccine rollout. Simulations with the agent-based model were conducted under the same conditions as the main analysis but assuming a 47% vaccine rollout (vaccine doses were tripled in simulation scenarios) that was prioritized to those who are ≥65 years old, living in nursing homes, or are medical workers, with additional doses distributed to those ≥18 years old. Results are presented as the number of infections or deaths averted relative to a base case of an aggregate of non-pharmaceutical interventions (102,946,388 cumulative infections and 338,222 cumulative deaths) based on monoclonal antibody supply from January 2021 of 300,000 doses/month (A) and 600,000 doses/month (B). The colored columns reflect distinct paradigms, with shading indicating different scenarios within the paradigm. The columns enclosed by broken lines additionally incorporate the use of rapid diagnostic tests.

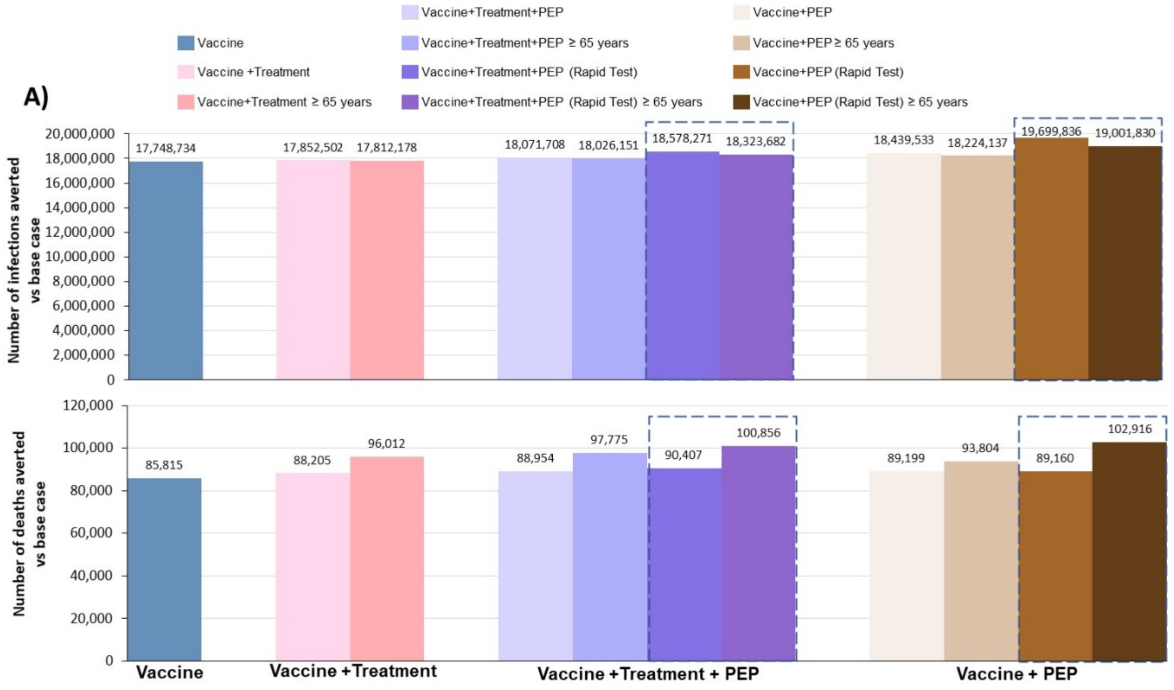

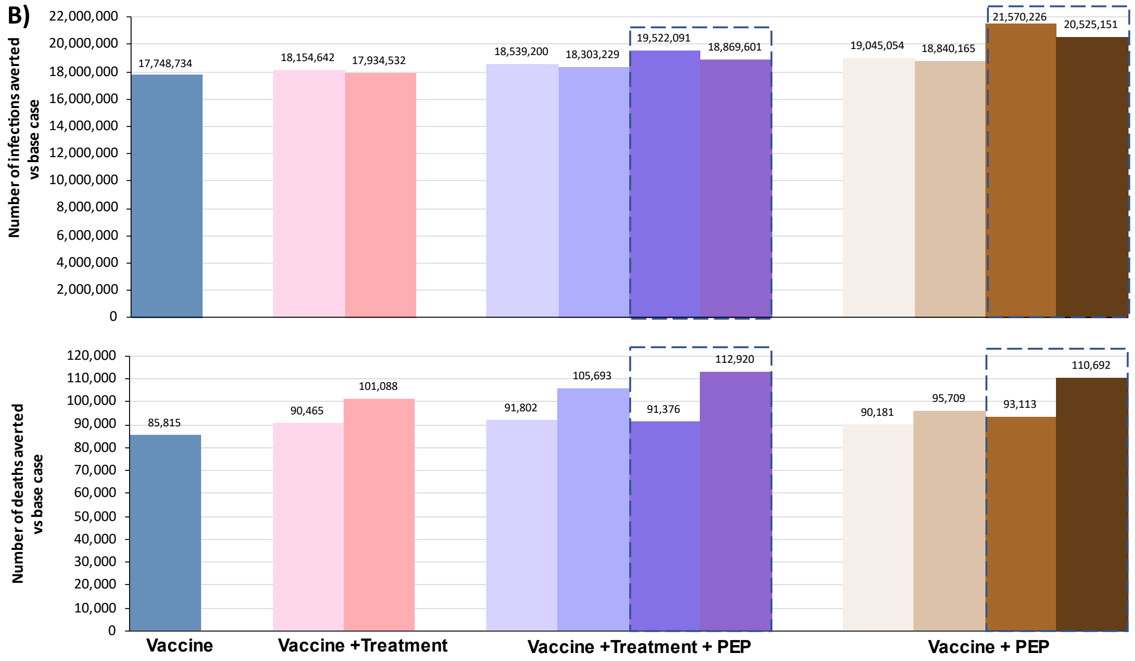
B)
Number of infections averted vs base case
22,000,000
20,000,000
18,000,000
16,000,000
14,000,000
12,000,000
10,000,000
8,000,000
6,000,000
4,000,000
2,000,000
0
17,748,734
18,154,642
17,934,532
18,539,200
18,303,229
19,522,091
18,869,601
19,045,054
18,840,165
21,570,226
20,525,151
Number of deaths averted vs base case
120,000
110,000
100,000
90,000
80,000
70,000
60,000
50,000
40,000
30,000
20,000
10,000
0
85,815
90,465
101,088
91,802
105,693
91,376
112,920
90,181
95,709
93,113
110,692
Vaccine
Vaccine +Treatment
Vaccine +Treatment + PEP
Vaccine + PEP

**Table S1** US Census Bureau tables used to derive agent-level characteristics to generate a population similar to that of the US.

| **US Census Table** | **Description** | **Source** |
|---|---|---|
| Table B01001 | Population by County<br><br>*County-level data* | US Census Bureau (2018). SEX BY AGE, 2013-2018 American Community Survey 5-year estimates. Retrieved from https://data.census.gov/cedsci/table?g=0100000US.050000&tid=ACSDT5Y2018.B01001&hidePreview=true |
| Table B14004 | Education by age for those ≥15 years old<br><br>*County-level data* | US Census Bureau (2018). SEX BY COLLEGE OR GRADUATE SCHOOL ENROLLMENT BY TYPE OF SCHOOL BY AGE FOR THE POPULATION 15 YEARS AND OVER, 2013–2018 American Community Survey 5-year estimates. Retrieved from https://data.census.gov/cedsci/table?g=0100000US.050000&tid=ACSDT5Y2018.B14004&hidePreview=true |
| Table B14005 | Education status and employment among those 16–19 years -old<br><br>*County-level data* | U.S. Census Bureau (2018). SEX BY SCHOOL ENROLLMENT BY EDUCATIONAL ATTAINMENT BY EMPLOYMENT STATUS FOR THE POPULATION 16–19 YEARS, 2013–2018 American Community Survey 5-year estimates. Retrieved from https://data.census.gov/cedsci/table?g=0100000US.050000&tid=ACSDT5Y2018.B14005&hidePreview=true |
| Table B23001 | Employment status among those ≥ 16 years old<br><br>*County-level data* | US Census Bureau (2018). SEX BY AGE BY EMPLOYMENT STATUS FOR THE POPULATION 16 YEARS AND OVER, 2013–2018 American Community Survey 5-year estimates. Retrieved from https://data.census.gov/cedsci/table?g=0100000US.050000&tid=ACSDT5Y2018.B23001&hidePreview=true |
| Table B24010 | Occupational status for employed civilians ≥ 16 years old<br><br>*County-level data* | U.S. Census Bureau (2018). SEX BY OCCUPATION FOR THE CIVILIAN EMPLOYED POPULATION 16 YEARS AND OVER, 2018 American Community Survey 1-year estimates. Retrieved from https://data.census.gov/cedsci/table?g=0100000US.050000&tid=ACSDT1Y2018.B24010&hidePreview=true |
| Table B24010 | Occupational status for employed civilians ≥16 years old<br><br>*State-level data* | U.S. Census Bureau (2018). SEX BY OCCUPATION FOR THE CIVILIAN EMPLOYED POPULATION 16 YEARS AND OVER, 2018 American Community Survey 1-year estimates. Retrieved from https://data.census.gov/cedsci/table?tid=ACSDT1Y2018.B24010&hidePreview=true |
| Table B08006 | Types of transportation used by workers<br><br>*County-level data* | U.S. Census Bureau (2018). SEX OF WORKERS BY MEANS OF TRANSPORTATION TO WORK, 2013-2018 American Community Survey 5-year estimates. Retrieved from https://data.census.gov/cedsci/table?g=0100000US.050000&tid=ACSDT5Y2018.B08006&hidePreview=true |
| Table B09021 | Living arrangements among adults ≥18 years old<br><br>*County-level data* | U.S. Census Bureau (2018). LIVING ARRANGEMENTS OF ADULTS 18 YEARS AND OVER BY AGE, 2013–2018 American Community Survey 5-year estimates. Retrieved from https://data.census.gov/cedsci/table?g=0100000US.050000&tid=ACSDT5Y2018.B09021&hidePreview=true |

| | | |
|---|---|---|
| Table B11016 | Household type by household size<br><br>*County-level data* | U.S. Census Bureau (2018). HOUSEHOLD TYPE BY HOUSEHOLD SIZE, 2013–2018 American Community Survey 5-year estimates. Retrieved from https://data.census.gov/cedsci/table?g=0100000US.050000&tid=ACSDT5Y2018.B11016&hidePreview=true |
| Table B23008 | Age of own children < 18 years and subfamilies by living arrangements<br><br>*County-level data* | U.S. Census Bureau (2018). AGE OF OWN CHILDREN UNDER 18 YEARS IN FAMILIES AND SUBFAMILIES BY LIVING ARRANGEMENTS BY EMPLOYMENT STATUS OF PARENTS, 2013–2018 American Community Survey 5-year estimates. Retrieved from https://data.census.gov/cedsci/table?g=0100000US.050000&tid=ACSDT5Y2018.B23008&hidePreview=true |
| Table P42 | Group quarters population by type of group quarters<br><br>*County-level data* | U.S. Census Bureau (2010). GROUP QUARTERS POPULATION BY GROUP QUARTERS TYPE, 2010 Decennial Summary File 1. Retrieved from https://data.census.gov/cedsci/table?g=0100000US.050000&tid=DECENNIALSF12010.P42&hidePreview=true |
| Table P42 | Group quarters population by type of group quarters<br><br>*State-level data* | U.S. Census Bureau (2010). GROUP QUARTERS POPULATION BY GROUP QUARTERS TYPE, 2010 Decennial Summary File 1. Retrieved from https://data.census.gov/cedsci/table?g=0100000US.04000.001&tid=DECENNIALSF12010.P42&hidePreview=true |
| Table B26101 | Age and sex population structure<br><br>*State-level data* | U.S. Census Bureau (2018). GROUP QUARTERS TYPE (3 TYPES) BY SEX BY AGE, 2013–2018 American Community Survey 5-year estimates. Retrieved from https://data.census.gov/cedsci/table?g=0100000US.04000.001&tid=ACSDT5Y2018.B26101&hidePreview=true |
| National School Enrollment | National school enrollment by type of school | U.S. Census Bureau (2018). Enrollment Status of the Population 3 Years Old and Over, by Sex, Age, Race, Hispanic Origin, Foreign Born, and Foreign-Born Parentage: October 2018, 2018 National School Enrollment. Retrieved from https://www2.census.gov/programs-surveys/demo/tables/school-enrollment/2018/2018-cps/tab01-01.xlsx |
| National Counties Gazetteer | | US Census Bureau (2019). Gaz Counties National, 2019 Gazetteer Files. Retrieved from https://www2.census.gov/geo/docs/maps-data/data/gazetteer/2019_Gazetteer/2019_Gaz_counties_national.zip |

**Table S2** Distribution of household composition in the US (source: US Census Bureau, 2018 American Community Survey).

| **Household size** | **Proportion (%)** |
|---|---|
| 1-person household | 27.8 |
| 2-person household | 33.9 |
| 3-person household | 15.7 |
| 4-person household | 13.0 |
| 5-person household | 6.0 |
| 6-person household | 2.3 |
| 7-person household | 1.3 |

**Table S3** Data sources for populating the movement module.

| Description | Source |
|---|---|
| Data used to calculate the age-based daily probability of staring a new trip and the probability of trip duration as shown in Table S4 and Table S5 | 1. HolidayTravel.qxp (dot.gov)<br>2. Cover.eps (workingcarsforworkingfamilies.org)<br>3. https://www.census.gov/data/tables/time-series/demo/popest/intercensal-2000-2010-national.html<br>4. TSA checkpoint travel numbers (current year(s) versus prior year/same weekday) \| Transportation Security Administration<br>5. https://www.arrivalist.com/daily-travel-index/ |
| Data used to calculate trip destination as shown in Fig. S7 | https://www.bts.gov/browse-statistical-products-and-data/surveys/american-travel-survey |

**Table S4** Age-based daily probability of starting a new trip.

| **Age (year)** | <25 | 25–64 | ≥65 |
|---|---|---|---|
| **Probability** | 0.0154 | 0.0266 | 0.0145 |

[a]The probabilities were reduced by 20% (average year-to-year reduction from June to December 2020) in the simulations to mimic the change of movement due to COVID-19 pandemic.

**Table S5** Probability of duration of each new trip.

| **Duration(days)** | 1 | 2–4 | 5–8 | 9–12 |
|---|---|---|---|---|
| **Probability** | 0.500 | 0.358 | 0.101 | 0.0407 |

**Table S6** Parameters calibrated and used in disease progression module as shown in Fig. S8.

| Parameter description | Values[a] |
|---|---|
| Probability of asymptomatic infection by age group | 0–9 = 0.456; 10–19 = 0.412; ≥20 = 0.4 |
| Probability of mild infection by age group | 0–9 = 0.73021; 10–19 = 0.77953; 20–29 = 0.81789; 30–39 = 0.84118; 40–49 = 0.84392; 50–59 = 0.82474; 60–69 = 0.78227; 70–79 = 0.71651; ≥80 = 0.63157 |
| Probability of hospitalization by age group | 0–9 = 0.001; 10–19 = 0.006; 20–29 = 0.015; 30–39 = 0.069; 40–49 = 0.219; 50–59 = 0.279; 60–69 = 0.37; 70-79 = 0.391; ≥80 = 0.379 |
| Probability of ICU needs by age group | 0–9 = 0.000386363; 10–19 = 0.000386363; 20–29 =0.00233248; 30–39 = 0.00233248; 40–49 = 0.00233248; 50–59 = 0.10425; 60–69 = 0.234135; 70–79 = 0.433395; ≥80 = 0.71129 |
| Probability of death in ICU by age group | 0–9 = 0.2970; 10–19 = 0.2970; 20–29 = 0.3465; 30–39 = 0.3465; 40–49 = 0.3780; 50–59 = 0.6840; 60–69 = 0.7740; 70–79 = 0.8550; ≥80 = 0.8910 |

[a]The initial values were from [11-13]. Those parameters were finally obtained from model calibration, thus they were suitable for our model and the target simulation scenarios. A cautious approach should be used when comparing those parameters with epidemiological data or applying them to other models.

**Table S7** Summary of main assumptions in simulation scenarios

| **Parameters** | | **Sources** |
|---|---|---|
| **Antiviral therapy** | | |
| Average time to treatment (day) of active treatment | 3 (SD = 1.4) days post symptom onset | Assumption |
| Average time to treatment (day) for post-exposure prophylaxis | 3 (SD = 1) days post infection | Assumption |
| Dose of active treatment | 1 dose (2400mg) | Clinical trial[8] |
| Dose of post-exposure prophylaxis | 0.5 dose (1200 mg) | Clinical trial[14] |
| Length of protection for prophylaxis | 30 days | Assumption |
| Efficacy of post-exposure prophylaxis when administered to susceptible agent (probability of being completely immune to infection after treatment) | 100%[a] | Assumption |
| Effectiveness of contact tracing (proportion of close contacts reached) | 100% for household members; 40% for colleagues and classmates | Assumption |
| Proportion of post-exposure prophylaxis administered to confirmed cases | 25% without rapid test; 100% with rapid test | Assumption |
| Sensitivity and specificity of rapid test (rapid diagnostics) | No assumption around test sensitivity or specificity (assume any test approved by FDA would have reasonable performance). | |
| Average time to rapid test (day) | Assumed that the rapid test was administered the same day as drug administered as post-exposure prophylaxis. | |
| Reduction of infectiousness | Modulated via viral load values, dependent on time of treatment initiation as described in "Antiviral treatment module" | |
| Reduction of disease progression | Modulated by reducing the probability of progressing to the following worse disease stage, consistent with clinical trial information As described in "Antiviral treatment module" | |
| **Disease progression** | | |
| The time from infection to symptom onset (day) | 5 | [5] |
| Duration of being exposed (E in Fig. S8) and not infectious (day) | 2 | [6] |
| Duration of being pre-symptomatic (Incu in Fig. S8) and infectious (day) | 3 | Time from infection to symptom onset – duration of being exposed and not infectious |
| Duration of being asymptomatic (A in Fig. S8) | 7 | Based on the simulated viral load values (100 |

| | | |
|---|---|---|
| | | control profiles as described in "Antiviral treatment module") multiplied by a factor of 75% |
| Duration of being symptomatic with mild symptoms (day), Mild in Fig. S8 | 6 | [15] |
| Duration of being symptomatic with severe symptoms before hospitalization (day), severe in Fig. S8 | 5 | [11] |
| Time needed to recover from severe symptoms if not hospitalized (day), Severe_rec in Fig. S8 | 2 | Assumed as equal to hospital stay before ICU |
| Duration of hospitalization if ICU not required (day) | 9 | [11] |
| Duration of hospitalization before critical care admission (day), Hosp in Fig. S8 | 2 | [11] |
| Time needed to recover from hospitalization (day), Hosp_rec in Fig. S8 | 7 | Hospitalization duration if ICU not required – duration of hospitalization before ICU |
| Duration of ICU stay (day), ICU in Fig. S8 | 10 | [15] |
| Probability of disease progression | Table S6 | Results obtained from model calibration |

[a]The basis for this assumption was an administrative assessment of 409 seronegative participants, which demonstrated 100% risk reduction in proportion of participants with symptomatic events with REGEN-COV. The primary analysis now available reports a risk reduction of 81%.[16]

**Table S8** Number of vaccine doses distributed each week in simulations.

| **Time period** | Week of Dec 14, 2020 | Week of Dec 21, 2020 | Week of Dec 28, 2020 | Week of Jan 4, 2021 | Week of Jan 11, 2021 | Week of Jan 18, 2021 | Week of Jan 25, 2021 | After week of Jan 25, 2021 |
|---|---|---|---|---|---|---|---|---|
| **Doses** | 1134552 | 1762667 | 2785164 | 5122554 | 135583 | 7688936 | 8060280 | Assume as 8060280 |

**Table S9** Data used to populate the vaccine module.

| **Description** | **Source** |
|---|---|
| Data used to calculate the vaccine doses as shown in Table S8. Data were managed on February 3, 2021. | https://covid.cdc.gov/covid-data-tracker/#vaccination-trends |

**Table S10** Treatment effects on reduction of overall infectiousness and infectious duration.

| **Day of treatment initiation post infection** | 1 | 2 | 3 | 4 | 5 | 6 | 7 | 8 | 9 | 10 |
|---|---|---|---|---|---|---|---|---|---|---|
| **Reduction of median infectiousness during the infectious days (AUC)** | Assume 90% | Assume 80% | 54% | 49% | 37% | 31% | 20% | 17% | 8% | 0% |
| **Reduction of median infectious duration (day)** | 88% | 63% | 50% | 50% | 38% | 38% | 25% | 25% | 13% | 0% |

**Table S11** Illustration of treatment effects on the reduction of disease progression.

| **Day of treatment initiation post infection** | 1 | 2 | 3 | 4 | 5 | 6 | 7 | 8 | 9 | 10 |
|---|---|---|---|---|---|---|---|---|---|---|
| **Reduction of the probability of disease progression** | 100% | 100% | 80% | 70% | 70% | 70% | 70% | 70% | 70% | 70% |

**Table S12** Infection fatality ratio (Source, https://www.cdc.gov/coronavirus/2019-ncov/hcp/planning-scenarios-archive/planning-ccenarios-2021-03-19.pdf).

| **Age (year)** | 0–19 | 20–49 | 50–69 | ≥70 |
|---|---|---|---|---|
| **Infection fatality ratio** | 0.00003 (0.00002, 0.0001) | 0.0002 (0.00007, 0.0003) | 0.005 (0.0025, 0.010) | 0.054 (0.028, 0.093) |

**Table S13** Data sources used to estimate the illness attack rate.

| **Description** | **Source** |
|---|---|
| Data used to estimate illness attack rate. | 1. Cumulative death reported by CDC https://data.cdc.gov/Case-Surveillance/United-States-COVID-19-Cases-and-Deaths-by-State-o/9mfq-cb36<br>2. The death distribution by age group used in the simulations (from CDC on 23 Nov 2020, https://covid.cdc.gov/covid-data-tracker/#demographics<br>3. Infection fatality ratio reported by CDC https://www.cdc.gov/coronavirus/2019-ncov/hcp/planning-scenarios-archive/planning-ccenarios-2021-03-19.pdf |

**Table S14** Illness attack rate by age calculated from the cumulative death, death distribution by age and infection fatality ratio by age.

| Date | Overall | Children (0–18) | Adults (19–64) | Adults (≥65) |
|---|---|---|---|---|
| June 1, 2020 | 9.5 % | 3.3 % | 13.4 % | 3.4 % |
| November 1, 2020 | 22.4 % | 7.8 % | 31.5 % | 8.0 % |

**Table S15** Results from model calibration, matching household secondary attack rates (Ptrans(t) = 0.102).

| **Data[17]** | | | |
|---|---|---|---|
| Overall | Children (0–19) | Adults (20–59) | Adults (≥60) |
| 17.2 % (14.1–20.6) | 6.4 % (2.8–12.2) | 18.5 % (14.4–23.2) | 28.0 % (19.1–38.2) |
| **Model simulated results (results from simulation of 20,000 independent households)** | | | |
| Overall | Children (0–19) | Adults (20–59) | Adults (≥60) |
| 16.0 % | 10.2 % | 18.4% | 22.6% |

**Table S16** Results from model calibration, matching the illness attack rate between June 1 and November 1, 2020.

| **Illness attack rate from June 1 to November 1 2020** | | | | |
|---|---|---|---|---|
| | **Overall** | **Children (0–18)** | **Adults (19–64)** | **Adults (≥65)** |
| Status on June 1 according to the simulation of 1 million agents | 9.5% | 3.3% | 13.4% | 3.4% |
| Status on November 1 according to the simulation of 1 million agents | 21. 1% | 7.5% | 30.0% | 8.5% |

**Table S17** Simulated sources of infection during model calibration.

| Source of infection | Share |
|---|---|
| Household (primary) | 36% |
| Workplace | 27% |
| School | 20% |
| Neighborhood | 11% |
| Community | 6% |
| Total | 100% |

**Table S18** Calibrated parameters $C_i$ (i.e., the probability of a sufficient contact for transmission during one time step) on a population of 1 million agents.

| | **Exposed** | | | |
|---|---|---|---|---|
| | **Child 0–4** | **Child 5–18** | **Adult 19–64** | **Adult ≥65** |
| Household | 0.1615944 | 0.1615944 | 0.3635874 | 0.4543635 |
| Nursing home | | | 0.001 | 0.001 |
| Student housing | | 0.001 | 0.001 | 0.000001 |
| Workplace[a] | | 0.0002921366 | 0.02741823 | 0.00001033734 |
| Kindergarten | 0.0006082194 | 0.0006082194 | | |
| Elementary school | | 0.0006082194 | | |
| Middle school | | 0.0035234794 | 0.003567523 | |
| High school | | 0.0042281738 | 0.004281026 | 0.00001756718 |
| University | | 0.0042441738 | 0.016600752 | 0.0001430807 |
| Neighborhood | 0.000066988 | 0.000066988 | 0.0007105086 | 0.0000001404329 |
| Community | 0.00000650113 | 0.00000650113 | 0.0000971232 | 0.00000012490605 |

[a]The values shown were assumed as default values for the office jobs. For client-facing jobs and medical jobs, we multiply the default values by a factor 2.17 and 7.59.[18], respectively. Note that, all those were included inside the model calibration

**Table S19** Calibrated infection fatality ratio.

| **Age (years)** | 0–19 | 20–49 | 50–69 | ≥70 |
|---|---|---|---|---|
| **Results according to simulation of 1 million agents** | 0.00005 | 0.0003 | 0.004 | 0.04 |

**Table S20** $P_{trans}(t)$ values obtained from model calibration.

| **Time** | Oct 26, 2020 | Nov 9, 2020 | Nov 23, 2020 | Dec 7, 2020 | Dec 21, 2020 | Jan 4, 2021 | Jan 18, 2021 | Feb 1, 2021 | Feb 15, 2021 | Mar 1, 2021[a] |
|---|---|---|---|---|---|---|---|---|---|---|
| **Ptrans(t) values** | 0.152814 | 0.133563 | 0.15067 | 0.206261 | 0.2174 | 0.245275 | 0.274179 | 0.282587 | 0.306112 | 0.320576 |

[a]This value was used for simulating the 2 weeks between March 1 and March 15, 2021. Since we used death data up to April 4, 2021 for calibration and the time lag between infection to death was 22 days in the model, the $P_{trans(t)}$ values that were needed for running simulations after March 15 were assumed to be 0.320576.

**Table S21** Data sources for longitudinal death.

| **Description** | **Source** |
|---|---|
| Longitudinal daily death reported by CDC | 1. https://data.cdc.gov/Case-Surveillance/United-States-COVID-19-Cases-and-Deaths-by-State-o/9mfq-cb36 |
| Longitudinal daily death reported by IHME | 2. https://covid19.healthdata.org/united-states-of-america?view=resource-use&tab=trend&resource=all_resources |

**Table S22** Dose allocation in post-exposure prophylaxis (PEP) as ratio of doses used for non-infected to infected individuals.

| **Scenarios in Figure 3 (A)** | **Ratio of doses used on non-infected: infected for PEP without rapid testing**[a] |
|---|---|
| Baseline + vaccine + PEP | 4.2 |
| Baseline + vaccine + PEP (≥ 65 years old) | 3.1 |
| Baseline +vaccine + treatment + PEP | 4.2 |
| Baseline + vaccine + treatment + PEP (≥65 years old) | 2.7 |

[a]These values were derived from simulations to illustrate how many doses were actually administered to susceptible agents when the drug was consumed as PEP without rapid testing.

**Table S23** Impact of vaccines on burden of infections and deaths during the aggressive pandemic phase (October 26, 2020 to April 4, 2021).

| **Efficacy after first dose** | **Days between 2 doses** | **Total number of vaccine doses** | **Proportion of population immunized (%)** | **Cumulative population infected over simulation (percent change)** | **Cumulative death over simulation (percent change)** |
|---|---|---|---|---|---|
| No pharmaceutical intervention | - | - | - | 102,946,388 | 338,221 |
| 52%[7] | 25 | 106,343,166 | 15.7 | 96,954,689 (-5.8%) | 295,217 (-12.7%) |
| | 60 | 106,370,393 | 16.3 | 96,611,411 (-6.2%) | 301,159 (-11.0%) |
| | 90 | 106,295,150 | 16.1 | 96,426,659 (-6.3%) | 301,805 (-10.8%) |

11. OpenABM-Covid19: Agent-based model for modelling the Covid-19 and Contact-Tracing. https://github.com/BDI-pathogens/OpenABM-

Covid19/blob/master/documentation/covid19.md. https://github.com/BDI-pathogens/OpenABM-Covid19/blob/master/documentation/covid19.md (accessed.
